# Coming Into Resistance: Clinically Important Antibiotic Resistance Genes in NICU vs. Healthy Infants

**DOI:** 10.64898/2026.09.23.26363796

**Authors:** Rachel E. Strength, Angelina Angelova, Emily Robbins, Shira Levy, Kathryn E. McCauley, Kevin M. Lloyd, Elisa B. Margolis, Andrew S. Burns, Shreni Mistry, Poorani Subramanian, Aimee Dassner, Christa Zerbe, Victor I. Band, Sivaranjani Namasivayam, Qing Chen, Esther Esadah, Katherine Rees, Mickayla Bacorn, Ruhika Prasad, Phoebe LaPoint, Hector N. Romero-Soto, John P. Dekker, Cynthia L. Sears, Craig Shapiro, Rana F. Hamdy, Joseph M. Campos, Suchitra K. Hourigan

## Abstract

**Importance:** Antibiotic-resistant infections in neonates often arise from the infant’s own colonizing bacteria harboring antibiotic resistance genes (ARG). This study evaluates ARG that impact antibiotic selection and drive resistance-related mortality, termed clinically important antibiotic resistance genes (CI-ARG).

**Objective:** To determine differences in gut CI-ARG abundance in children from birth through 3 years old, stratified by neonatal intensive care unit (NICU) admission and antibiotic (abx) exposure in the first two months

**Design:** Cohort study (sequencing, analysis completed 2025-2026) using shotgun metagenomic sequencing of stool samples from infants in two studies: “Neonatal Intestinal Microbiome” (+NICU groups, 2015-2022), “The First 1000 Days of Life and Beyond” (−NICU groups, 2018-2021). Infants grouped into four cohorts: +NICU/+abx, +NICU/−abx, −NICU/−abx, and −NICU/+abx. Prespecified CI-ARG abundance compared longitudinally among groups.

**Setting:** NICU, well-baby nursery, Inova Fairfax Hospital, Virginia

**Participants:** +NICU criteria: admission to the NICU, anticipated length of stay (LOS) of at least five days, maternal age ≥18 years. −NICU: parents with good general health and age ≥18 years enrolled prior to infant birth. Stool samples: adequate stool processing volume, at least one stool sample from before one month and after one year of age, 7-day minimum between serial samples.

**Exposures:** NICU versus well-baby nursery; antibiotics versus none in first two months of corrected age

**Main Outcomes and Measures:** Predefined primary outcome-longitudinal CI-ARG abundance by group, compared using linear mixed effects models with adjustment for multiple comparisons (Benjamini-Hochberg procedure). Predefined secondary outcomes-differences in overall microbiota and resistome composition.

**Results:** 1000 serial stool samples, 179 infants (birth-39 months). Longitudinal abundance significantly differed among cohorts for 11 CI-ARG (p<0.05); often higher in the +NICU groups in early life, with or without antibiotic exposure. Abundance of ARG conferring resistance to aminoglycosides, macrolides, and fluoroquinolones later decreased below −NICU levels. *mecA*, which often drives methicillin resistance in *Staphylococcus*, was detected only in +NICU infants during premature period.

**Conclusions:** This longitudinal study demonstrates significant longitudinal CI-ARG differences in children grouped by early-life NICU status and/or antibiotic exposure. Future studies should explore the mechanisms driving these CI-ARG changes and evaluate their impact on infection risk and community spread.

**Key Points:** *Question:* How do clinically important antibiotic resistance genes (CI-ARG) differ longitudinally among infants up to 3 years old based on NICU admission and antibiotic exposure in the first two months of life?

*Findings:* In this cohort study of 179 infants through 39 months of age, longitudinal abundance was significantly different for 11 prespecified CI-ARG, often higher in the NICU groups in early life, with or without antibiotic exposure.

*Meaning:* Children with a history of NICU hospitalization, whether due to the NICU environment itself, prematurity, or other factors, may have increased CI-ARG burden early in life, irrespective of direct antibiotic exposure.

## Introduction

In 2021, over 840,000 deaths in children under five years old were associated with antimicrobial resistance (AMR) globally.^1^ Bacteria commonly found in the gut, such as *Escherichia coli*, accounted for over 80% of these deaths.^1^ The gut microbiome has been proposed as a reservoir for AMR spread, both within an individual and between communities.^2,3^ Because early life is a critical window when commonplace exposures can have lasting effects on microbiome composition and host health, understanding how antibiotic resistance gene (ARG) abundance evolves during this period is essential.^4^

Antibiotic resistance genes are detectable in the infant gut from the first days of life, regardless of antibiotic exposure.^5^ The infant gut resistome comprises all ARG in the gastrointestinal tract and is affected by numerous factors: antibiotic use, breastfeeding, mode of delivery, gestational age, and microbiota composition.^6^ Some of these factors may be especially important in the neonatal intensive care unit (NICU) setting, where interventions like parenteral nutrition and surgery further influence the microbiota.^7^ Children under two years old have significantly higher ARG abundance than adults living in a similar environment, even more than their own mother in the first six months.^8,9^ This elevated ARG burden and the inherent instability of the nascent microbiome may increase vulnerability to colonization by antibiotic-resistant organisms.^6,10^

While many ARG can give rise to AMR, certain genes commonly cause resistance to antibiotics critical for healthcare infrastructure, here termed clinically important antibiotic resistance genes (CI-ARG). For instance, *mecA* often drives beta-lactam resistance in *Staphylococcus* species, most notably in methicillin-resistant *S. aureus*.^11^ Prior studies have examined the infant gut resistome broadly without focusing on these genes specifically. The risks of CI-ARG are particularly relevant in NICU infants, who face frequent empiric antibiotic exposure and elevated infection risk from both commensals and pathogens.

While previous research has contributed to our understanding of AMR in infants, considerable knowledge gaps remain. Only one study has compared the gut resistomes of preterm versus term infants for more than one year, finding persistent metagenomic signatures of antibiotic therapy through 21 months in preterm infants with higher antibiotic exposure.^12^ There have been no studies comparing CI-ARG changes over time in NICU infants versus healthy infants with or without early-life antibiotic exposure. To address these underexplored areas, we compared our primary outcome of CI-ARG abundance, in addition to the secondary outcomes of overall resistome and microbiota composition, in NICU infants and healthy infants in a longitudinal cohort study over 39 months of corrected age (i.e. age corrected for gestation), stratified by NICU admission and early-life antibiotic use. We hypothesized that infants with NICU and antibiotic exposure would have higher CI-ARG abundance over time compared to those without these exposures.

## Methods

Neonates at Inova Fairfax Hospital in Virginia were enrolled in two temporally overlapping, longitudinal, prospective cohort studies: “Neonatal Intestinal Microbiome: Impact on Infant and Early Childhood Health and Disease” (+NICU cohort, 2015-2022) and “The First 1000 Days of Life and Beyond” (−NICU cohort, 2018-2021).^13,14^ The +NICU cohort inclusion criteria were NICU admission, anticipated length of stay (LOS) of at least five days, and maternal age ≥18 years. For the −NICU cohort in the well-baby nursery, the inclusion criteria were parental good general health and age ≥18 years.

Serial stool samples were collected from the infants while in the hospital and post-discharge. Every sample meeting the following criteria was included: adequate stool processing volume, at least one stool sample from before one month and after one year of age (for exceptions, see eMethods), and a minimum of seven days between serial samples. All samples underwent shotgun metagenomic sequencing on the NovaSeq platform (Illumina) with a sequencing depth goal of 25-30 million reads per sample.

Age refers to corrected age, i.e. day of life (DOL) adjusted for gestational age at birth and zeroed at expected full-term gestation (40 weeks).

To study CI-ARG in these children, we divided them into four cohorts based on systemic antibiotic (abx) use in the first two months of age and NICU admission after birth:

- +NICU/+abx (n=74)
- +NICU/−abx (n=54)
- −NICU/+abx (n=6; non-NICU neonates are often healthy)
- −NICU/−abx (n=45)

NICU admission could not be completely disentangled from confounders like gestational age and medical complexity, as premature, ill infants are preferentially admitted to the NICU. We thus chose not to adjust for gestational age, instead using corrected age to align +NICU and −NICU infants. The term “+NICU” denotes this broader collection of factors associated with NICU admission rather than NICU exposure alone.

CI-ARG were defined a priori. Analysis techniques included linear mixed effects models adjusted for longitudinal repeated measures and principal coordinates analysis (PCoA) of Bray-Curtis dissimilarity matrices. Taxonomic dynamics and CI-ARG comparisons by NICU and antibiotic strata were analyzed post-hoc for exploratory purposes; linkages between taxa and CI-ARG were computationally inferred from correlations in abundance patterns. Age binning was used for visualization (bin limits: eTable 1, sample distribution: eTable 2). Each analysis was adjusted for potential confounders, including infant sex, self-reported maternal race and ethnicity, delivery mode, intrapartum antibiotics, antibiotics during pregnancy, and infant antibiotic use since last sample (after two months of age, binary variable). See the eMethods for the full methodology.

## Results

The analysis included 179 infants with 1039 serial stool samples over the first 39 months of age. The groups were similar in terms of infant sex, maternal race and ethnicity, and antibiotic use after two months (Table 1). As expected, +NICU infants were more often delivered by Cesarean section, born at younger gestational ages, had more maternal antibiotic exposure, and had longer hospital LOS. Most infants had early-life breastmilk exposure, though inconsistent questionnaire responses precluded adjustment for diet per sample.

**Table 1:** Participant demographics. Bolded p-values indicate significant differences among the four study groups (p<0.05). P-values were not calculated for count variables like participant and sample numbers. Abx is an abbreviation for antibiotics, NA is an abbreviation for not applicable.

|  |  | +NICU |  | -NICU |  |  |
| --- | --- | --- | --- | --- | --- | --- |
|  |  | +Abx | -Abx | +Abx | -Abx | p-values |
| # Subjects |  | 74 | 54 | 6 | 45 | NA |
| # Samples |  | 506 | 330 | 24 | 179 | NA |
| Vaginal delivery |  | 19 (26%) | 7 (13%) | 2 (33%) | 28 (62%) | **<0.001** |
| Female sex |  | 27 (36%) | 25 (46%) | 3 (50%) | 22 (49%) | 0.52 |
| Maternal race | White | 37 (50%) | 28 (52%) | 5 (83%) | 31 (69%) | 0.41 |
|  | Black | 11 (15%) | 6 (11%) | 0 (0%) | 5 (11%) |  |
|  | Asian | 10 (14%) | 11 (20%) | 0 (0%) | 5 (11%) |  |
|  | Other | 16 (22%) | 9 (17%) | 1 (17%) | 4 (9%) |  |
| Maternal ethnicity | Hispanic/Latino | 9 (12%) | 4 (7%) | 0% | 3 (7%) | 0.13 |
| Gestational age (weeks) |  | Median: 30<br>IQR: 27-34 | Median: 33<br>IQR: 31-34 | Median: 39<br>IQR: 39-39 | Median: 39<br>IQR: 39-40 | **<0.001** |
| Length of stay (days) |  | IQR: 25.5-93.5<br>Median: 50 | IQR: 17-42.3<br>Median: 26 | IQR: 2-2.75<br>Median: 2 | IQR: 2-2<br>Median: 2 | **<0.001** |
| Maternal abx | Pregnancy | 25 (34%) | 9 (17%) | 1 (17%) | 6 (13%) | **0.033** |
|  | Intrapartum | 69 (93%) | 53 (98%) | 4 (67%) | 22 (49%) | **<0.001** |
| Antibiotics after two months corrected age |  | 41 (63%)<sup>a</sup> | 23 (49%)<sup>a</sup> | 4 (67%) | 17 (38%) | 0.052 |
| Any breastmilk exposure |  | 66 (89%) | 44 (81%) | 2 (33%)<br>3 unknown | 36 (80%)<br>1 unknown | **0.007** |

<sup>a</sup> Note different denominators due to sample availability (see eMethods)

Ampicillin and gentamicin were the most common antibiotics in the +NICU/+abx cohort in the first two months, while ampicillin, gentamicin, and ceftazidime were most common in the −NICU/+abx cohort (Table 2). In both cohorts, sepsis rule-out was the most frequent indication.

**Table 2:** Participant antibiotic use per cohort. Number of infants in each +abx cohort with exposure to a listed antibiotic in the first two months of corrected age. The reasons for antibiotic use are also included. Note that there may have been multiple reasons for antibiotic use in an infant, so the numbers do not sum to the total number of infants per group. The “Other” category includes perioperative prophylactic antibiotics and prophylaxis for conditions like gastroschisis.

| **Antibiotic** | **Number of -NICU/+abx infants with any abx exposure before 2 months corrected age** | **Number of +NICU/+abx infants with any abx exposure before 2 months corrected age** |
| --- | --- | --- |
| Amoxicillin | 1 | 1 |
| Ampicillin | 4 | 70 |
| Cefazolin | 0 | 8 |
| Cefotaxime | 0 | 1 |
| Cefoxitin | 0 | 1 |
| Ceftazidime | 2 | 8 |
| Cefuroxime | 0 | 1 |
| Cephalexin | 1 | 1 |
| Gentamicin | 2 | 70 |
| Metronidazole | 0 | 1 |
| Nafcillin | 0 | 1 |
| Piperacillin-tazobactam | 0 | 6 |
| Vancomycin | 0 | 15 |
| **Reason for Antibiotic** | **Number of -NICU/+abx infants given abx for that reason** | **Number of +NICU/+abx infants given abx for that reason** |
| Sepsis rule-out | 4 | 68 |
| Skin/soft tissue infection | 0 | 7 |
| Necrotizing enterocolitis | 0 | 3 |
| Lower respiratory tract infection | 0 | 3 |
| Urinary tract infection | 1 | 1 |
| Bacteremia (CoNS) | 0 | 1 |
| Central nervous system infection | 0 | 1 |
| Osteomyelitis | 0 | 1 |
| Phlebitis | 0 | 1 |

| Reason for Antibiotic<br>(continued) | Number of -NICU/+abx infants<br>given abx for that reason | Number of +NICU/+abx infants<br>given abx for that reason |
| --- | --- | --- |
| Otitis media | 1 | 0 |
| Other | 0 | 15 |

For alpha diversity, resistome Shannon indices differed significantly across the four cohorts despite similar richness (Figure 1a, p<0.001, Figure 1c, p=0.42, respectively). The microbiota alpha diversity increased over time in all cohorts, with significant differences in Shannon index but not richness (Figure 1b, p<0.001, Figure 1d, p=0.65, respectively).

**Figures 1a-1d:**
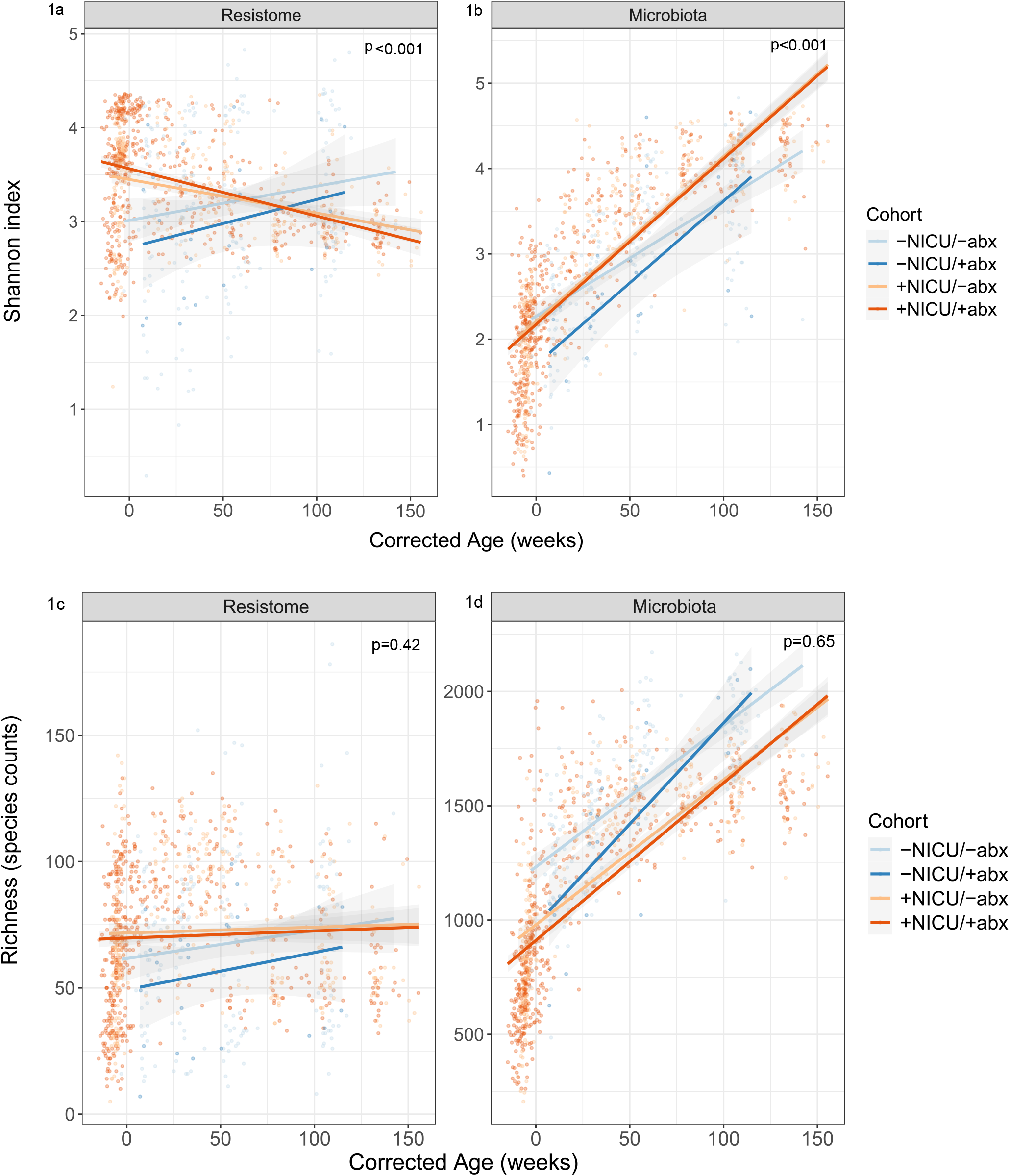
Linear mixed effect model plots of alpha diversity over time between cohorts. Linear mixed effects models were used to calculate longitudinal trajectories of the variable of interest (Shannon index and richness of the overall resistome or microbiota) over weeks of corrected age. Cohorts are denoted by color. Each point on the graphs represents the Shannon index or richness (species counts) of the resistome or microbiota of one sample. P-values were corrected for all identified confounders. **(1a) Resistome Shannon index. (1b) Microbiota Shannon index. (1c) Resistome richness. (1d) Microbiota richness**.

Examining beta diversity, the overall resistome composition was significantly different among cohorts (Figure 2a, PCo1 p<0.001, PCo2 p=0.002) and reflected in the overall microbiota composition (Figure 2b; PCo1 and PCo2 p<0.001). Additionally, when comparing across cohorts within each age bin post hoc, there were significant differences in both the resistome and microbiota in the 2-24-month age bins, where all cohorts had representative samples (eFigures 1a and 1b). None of the most abundant individual ARG overall were CI-ARG (eFigure 2). No significant gene-depth discrepancies suggestive of significant cohort-based or between-study differences were detected (eMethods, eFigure 3).

**Figures 2a-2b:**
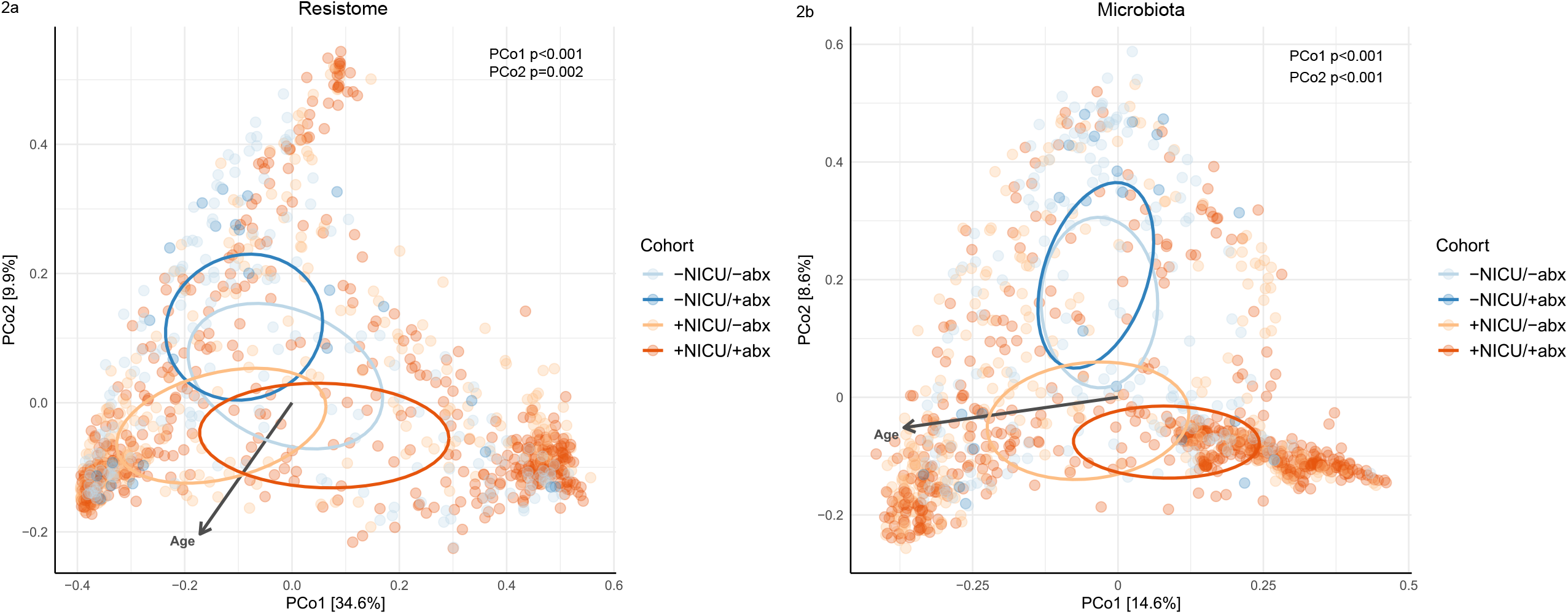
Principal coordinates analysis of beta diversity between cohorts, as measured by the Bray-Curtis dissimilarity metric. P-values produced from linear mixed effects models based on cohort were corrected for all identified confounders. Ellipsoids at 20% confidence regions were used to denote the centroid tendency of each group’s cluster. The gray arrow represents a fitted environmental vector of corrected age as a continuous variable, projected onto the PCoA space. One random sample per participant per age bin was used for analysis of that participant to prevent skewing of data (see eMethods). Cohorts are denoted by color. **(A) PCoA of resistomes by cohort. (B) PCoA of microbiota by cohort**.

Longitudinal CI-ARG abundance was compared across the four cohorts (CI-ARG list-eTable 3). CI-ARG abundance differed among the four cohorts for 11 CI-ARG (Figure 3). However, the two +NICU groups were often similar regardless of antibiotic use, as were the two −NICU groups.

**Figure 3:**
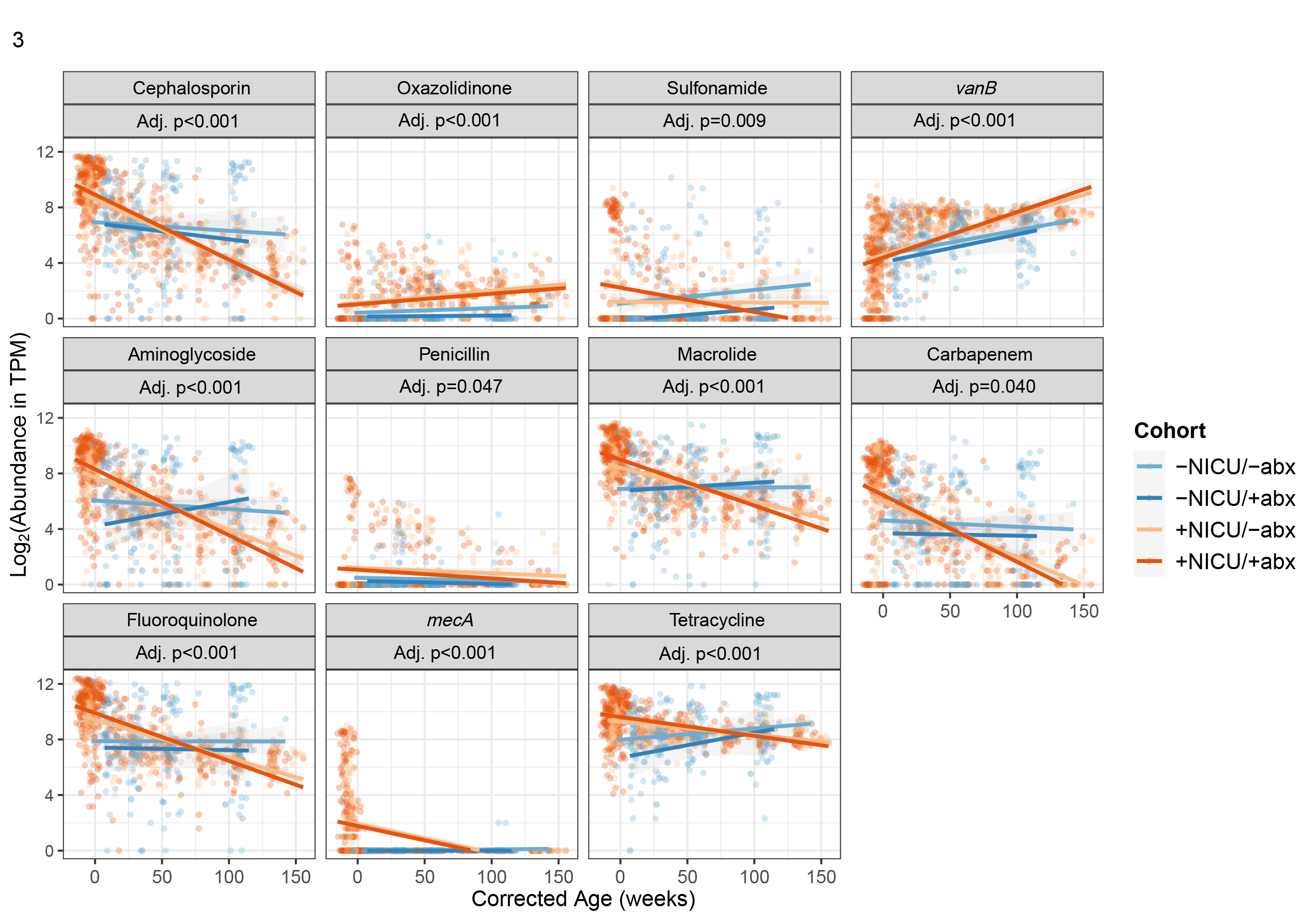
Linear mixed effects models of CI-ARG abundance over time by cohort. Only CI-ARG with statistically significant differences after Benjamini-Hochberg correction (false discovery rate < 0.05) with adjusted p-values between cohorts are shown. Each plot displays CI-ARG log_2_(abundance in transcripts per million, TPM) on the y-axis and corrected age in weeks on the x-axis. Each point represents the abundance of the listed CI-ARG in one sample. Cohorts are again noted by color, and p-values are corrected for all identified confounders.

As an exploratory analysis, we calculated nominal p-values for each of the prior figures to further investigate our findings that ARG changes appear more closely associated with NICU status than antibiotic exposure. Within each NICU stratum, antibiotic exposure was not associated with differences in resistome alpha diversity (eTable 4, p≥0.56), while all +NICU vs. −NICU comparisons were (p<0.001). Beta diversity analysis showed similar findings, (eTable 5, +abx vs. −abx p≥0.69, +NICU vs. −NICU p≤0.001).

Overall, NICU status was usually associated with differences in CI-ARG abundance (eTable 6); antibiotic exposure showed little evidence of a generalized association (eTable 7).

There were four observed patterns of variation across CI-ARG profiles in post hoc analysis.

In Pattern 1, the +NICU CI-ARG abundance was higher than the −NICU CI-ARG abundance, even when accounting for antibiotic exposure. This pattern was most consistent with our original hypothesis and was most evident in the abundance of *vanB* over time (Figure 3). We showed participant-level changes in resistance gene abundance along with timing of antibiotics for improved visualization (eFigure 4a).

Two of the top 25 taxa contributed significantly different amounts of *vanB* across cohorts, *Fusicatenibacter saccharivorans* and *Clostridium perfringens* (eFigure 4b). The taxon-specific contributions to *vanB* abundance (eFigure 4c) did not consistently match the overall *vanB* pattern (Figure 3). For example, inferred *C. perfringens vanB* carriage decreased in the +NICU groups while total *vanB* abundance increased. To explore this, we calculated the overall abundance trajectory of each differential taxon regardless of *vanB* carriage (eFigure 4d) and compared it to *vanB*-carrying subsets (eFigure 4c). In the +NICU cohorts, the decline in *C. perfringens vanB* carriage coincided with a decrease in its overall abundance, while *F. saccharivorans* demonstrated increased overall taxon abundance alongside increased *vanB* carriage (eFigures 4c-4d).

In Pattern 2, the +NICU CI-ARG abundance was high in early life, prior to the −NICU groups’ birth, then decreased to below the −NICU CI-ARG abundance. This pattern applied to most significantly different CI-ARG, including aminoglycoside resistance genes (hereafter, aminoglycoside ARG; Figure 3, eFigure 5a). This early-life aminoglycoside ARG elevation roughly coincides with gentamicin (an aminoglycoside) use in the NICU setting. *E. coli, Klebsiella pneumoniae*, and *Klebsiella michiganensis* were the top taxa inferred to carry aminoglycoside ARG, and the differentially contributing taxa between cohorts were *E. coli, Staphylococcus caprae*, and *Enterococcus gallinarum* (eFigure 5b). Despite large decreases in *E. coli* ARG carriage in the +NICU groups (eFigure 5c), overall *E. coli* taxon abundance decreased only slightly (eFigure 5d). *S. caprae* and *E. gallinarum* ARG and taxa abundances decreased considerably over time in the +NICU groups (eFigures 5c, 5d), similar to the +NICU decrease in aminoglycoside ARG abundance (Figure 3).

In Pattern 3, the +NICU CI-ARG abundance was higher than the −NICU read abundance in early life, then decreased to near zero (e.g. *mecA*). The vast majority of *mecA* was detected in +NICU infants prior to zero months (Figure 3, eFigure 6a). The main taxa putatively contributing to *mecA* abundance were non-aureus staphylococci (eFigure 6b). The abundance of both *mecA* and these species decreases over time (eFigures 6c, 6d), in concert with Figure 3’s pattern.

In Pattern 4, there were no significant differences in longitudinal CI-ARG abundance among the four groups. Some CI-ARG were not significantly different among groups (eTable 3). Many individual resistance genes were not detected, including *mecC* and genes producing carbapenemases such as VIM, IMP, KPC, and NDM.

Of note, ARG for oxazolidinones and penicillins followed Pattern 1; ARG for cephalosporins, macrolides, carbapenems, fluoroquinolones, tetracyclines, and sulfonamides followed Pattern 2 (Figure 3, eFigures 7-14). Participant-level abundance plots for all detected, non-significant CI-ARG are shown in eFigures 15-24.

## Discussion

To our knowledge, this is the first study comparing longitudinal CI-ARG trajectories with or without NICU and/or early-life antibiotic exposure. Conventional clinical thinking holds that antibiotic use results in more AMR. These results challenge that assumption somewhat, with longitudinal ARG changes in this population being complex and seemingly more dependent on NICU status than antibiotics in the long term. Contrary to our hypothesis that the +NICU/+abx cohort would have higher gut CI-ARG abundance longitudinally, many of those children had lower CI-ARG abundance than the −NICU cohorts at three years of age. Our study suggests that CI-ARG may follow relatively distinct temporal trajectories depending on NICU status, regardless of antibiotic use in early life.

There were significant differences in resistome Shannon indices over time. In the +NICU cohorts, resistome Shannon diversity declined longitudinally, contrasting the corresponding increase in microbiota diversity of those same groups. This could mean that, despite increases in the number of taxa in the +NICU cohorts, newly added bacteria may not be contributing different ARG (e.g. due to strain replacement, ARG loss, or colonization by ARG-poor taxa). These data align with other studies showing that the microbiota and resistome do not necessarily change in tandem.^12,15^ Resistome and microbiota beta diversity were significantly different among the four cohorts, consistent with prior research.^12^

We analyzed CI-ARG to capture longitudinal resistance patterns most applicable to clinical practice. Because ARG may be plasmid- or chromosome-associated, and the infant gut microbiome is particularly prone to horizontal gene transfer, we included all CI-ARG rather than restricting analyses to plasmid-based genes.^16^ This approach reflects the limitations of routine microbiologic ARG testing, which often does not distinguish gene location or bacterial host. We were surprised that antibiotic exposure in early life did not seem to have a stronger association in post hoc analysis. This could be because the infants’ antibiotic exposure was typically quite brief, frequently only 48 hours. Furthermore, some studies have shown that antibiotic exposure in the first week of life is less disruptive to the microbiome than later exposure, suggesting that the early-life microbiome’s dynamic nature may mitigate ARG persistence.^17^

The predominance of commensal carriers (e.g. *vanB* carriers) suggests that the previously described resistance trajectories could reflect broader community dynamics rather than pathogen-driven selection. The infant gut microbiome has been shown to have higher ARG and mobile genetic element carriage than adults, implying increased ARG transmission potential within and between individuals.^18,19^

While infant data remain sparse, research in adult populations has shown that hospital exposure, even without antibiotic use, changes the microbiota significantly.^20^ Our post-hoc exploratory analyses suggested that NICU admission (or NICU-associated factors like prematurity) had a stronger association with CI-ARG abundance than antibiotic exposure. Several mechanisms may explain this finding. The NICU microbiome is characterized by disrupted community structure, reducing ecological competition and allowing ARG-carrying strains to establish and persist despite an ARG’s fitness cost.^21^ As the microbiome matures after discharge, competitive pressure from incoming commensals may selectively displace low-fitness, ARG-carrying strains—a dynamic consistent with the Pattern 2 finding that +NICU CI-ARG abundance fell below −NICU levels in later life. Additional contributing factors may include horizontal gene transfer facilitated by the dense NICU microbial environment, selective pressure from environmental antibiotic residue, and founder effects from early microbial colonization in a low-diversity community.

While many individual CI-ARG are of low abundance compared to the most common ARG, they remain clinically relevant. The significance of these low-abundance ARG is best demonstrated by the clinical utility of highly sensitive, PCR-based methods of ARG surveillance and evidence that patients colonized with resistant organisms face increased infection risk regardless of abundance.^22^ In patient care, resistome composition often matters more than size or diversity. For example, even small populations of extended-spectrum-beta-lactamase producers in critically ill patients would likely be more concerning than larger, more diverse resistomes containing less harmful ARG.

Examining CI-ARG patterns more closely, in Pattern 1, +NICU CI-ARG abundance was higher than −NICU CI-ARG abundance, most consistent with our initial hypothesis (e.g. *vanB*). Computationally inferred taxon-level patterns did not consistently mirror cohort-level abundance, suggesting that overall *vanB* dynamics may reflect small, cumulative contributions from multiple taxa rather than large individual taxon shifts.

In Pattern 2, +NICU CI-ARG abundance was higher early in life, then decreased to below the −NICU CI-ARG abundance. The commonality of Pattern 2 across many CI-ARG may indicate ARG co-occurrence, as shown elsewhere.^23^ In the +NICU groups, the decline in aminoglycoside ARG abundance was linked to reduced ARG-carrying *E. coli*. However, the magnitude of +NICU ARG decline exceeded the decrease in overall *E. coli* abundance, suggesting ARG loss or strain replacement rather than whole-taxon loss. In contrast, decreases in *S. caprae* and *E. gallinarum* parallelled reductions in taxon-associated ARG carriage, suggesting possible taxon loss.

Although aminoglycosides were frequently administered in this study, aminoglycoside ARG trajectories were similar across NICU strata regardless of antibiotic exposure.

Notably, CI-ARG abundance in +NICU groups became lower than in −NICU groups later in life, again potentially due to ARG fitness cost.^24^ Although the higher +NICU ARG abundance is not sustained long-term, the early-life increase may still be relevant, given that preterm infants face elevated mortality risk in the first year of life.^25^ Prior research has shown that many ARG increase over the first year of life in healthy infants, including aminoglycoside ARG.^26^ Our study expands these data.

In Pattern 3 with *mecA*, +NICU CI-ARG abundance began higher than the −NICU groups, then decreased to near zero. Early-life *Staphylococcus* population collapse has been demonstrated previously; this could be the basis for *mecA* decline.^27^

In Pattern 4, there were no significant differences in longitudinal CI-ARG abundance. Many classically plasmid-based ARG (e.g. *bla*_KPC_) were not detected in this study, possibly due to diverse plasmid sizes, low copy number (a challenge in shotgun metagenomic sequencing), or CI-ARG acquisition later in life.^28^

This study has several limitations. Breastfeeding data lacked sample-level granularity, limiting our ability to control for feeding modality, though most children in this study had early-life breastmilk exposure. Next, there were differences in sampling frequency and cohort size between groups; these were accounted for during analysis. Despite the relatively small size of the −NICU/+abx group, its sample completeness and concordance with the −NICU/-abx group’s taxonomic and resistome dynamics support its retention in this analysis. Additionally, metagenomic detection of ARG does not confirm phenotypic resistance or bacterial viability, point mutations conferring resistance may not be well-detected with shotgun sequencing, and ARG identification is constrained by database availability of documented sequences. In evaluating some CI-ARG by antibiotic drug class, there is also higher risk of detecting ARG with lower clinical relevance. Finally, antibiotic exposure was a binary variable in this study and did not account for spectrum or duration, and the low number of infections in any group precluded meaningful analysis of the relationship between CI-ARG and clinical outcomes.

Future research should track CI-ARG acquisition and transmission within and between infants in longer, multi-environment studies to better understand ARG spread.

Furthermore, the relationship between higher CI-ARG carriage and antibiotic-resistant infections remains unestablished in infants, though it has been shown in other groups.^29^ Regardless of infection risk, carriage itself may be a source of community AMR spread.^30^

Infant gut CI-ARG variations over time are complex and not necessarily in line with clinical doctrine that antibiotics increase resistance uniformly. Resistance gene carriage may be significantly associated with environmental factors aside from antibiotic treatment, such as NICU status. While further research is needed, this study provides insight into longitudinal CI-ARG changes during a critical period of microbiome development.

## Supporting information

Online-Only Supplement

## Acknowledgments

The content is solely the responsibility of the authors and does not necessarily represent the official views of the National Institutes of Health. The authors thank the participants and their families for their dedication to this project as well as Kristy and Roger Crombie for their generous philanthropic donation toward this project in loving memory of their daughter Anna Charlotte. The authors also thank the NIH Intramural Sequencing Center (NISC) for assistance with sequencing. This study used the Office of Cyber Infrastructure and Computational Biology (OCICB) High Performance Computing (HPC) cluster at the National Institute of Allergy and Infectious Diseases (NIAID), Bethesda, MD. The authors also thank Dr. Andrea Hahn for her valuable input on this paper including feedback on the study analysis and editing.

The following authors are or were employed by the National Institutes of Health (who funded this study) at the time of their work: Rachel E. Strength, Angelina Angelova, Emily Robbins, Shira Levy, Kathryn E. McCauley, Andrew S. Burns, Shreni Mistry, Poorani Subramanian, Christa Zerbe, Victor I. Band, Sivaranjani Namasivayam, Qing Chen, Katherine Rees, Mickayla Bacorn, Ruhika Prasad, Phoebe LaPoint, Hector N. Romero-Soto, John P. Dekker, Suchitra K. Hourigan. Rachel E. Strength owns small, long-term stock holdings (<$1500 total) in Vertex Pharmaceuticals, Merck, Thermo Fisher Scientific, and Johnson & Johnson. These are passive personal investments and did not influence the study’s conduct or findings. Aimee Dassner, PharmD received support for conference travel from her current employer Thermo Fisher Scientific; she was previously employed at Children’s National Hospital for the majority of the work completed in this paper. Victor I. Band has US patent US12509714B2, “Multiple heteroresistance to guide combination antibiotic regimens.” All other authors have declared that they have no conflicts of interest.

Rachel Strength and Suchitra Hourigan had full access to all the data in the study and take responsibility for the integrity of the data and the accuracy of the data analysis. The following authors have accessed the data, verified the data, and were responsible for the data analysis: RES, AA, ER, SL, KEM, PS, SKH.

## Ethics approval and consent to participate

All participants provided signed informed consent prior to collection and storage of samples. This study was Institutional Review Board approved (WCG IRB 1300205 and WCG protocol 20120204). Consent forms included language authorizing publication of findings. Compensation was given to +NICU participants but not to −NICU participants per the funding structures of each study.

## Data Sharing

Data are publicly available from the Sequence Read Archive for the well-baby nursery cohort under the accession PRJNA988496, and for the NICU cohort under the accession PRJNA1280936. Deidentified participant data, the data dictionary, the study protocol, the statistical analysis plan, and the informed consent form will be made available upon reasonable request beginning at the time of publication. Data requests may be submitted to the senior author Suchitra Hourigan, MD at. Requests will be reviewed and approved by the corresponding author or the institutional data access committee. Data will be made available to any qualified researcher for any legitimate research purpose, subject to completion of a signed data access agreement. Data will be shared without investigator support unless otherwise agreed upon at the time of the request.

This paper does not report original code.

## Author Contributions

Conceptualization: RES, ER, SKH

Methodology: RES, AA, ER, SL, KEM, KML, EBM, AD, CZ, VIB, SN, QC, EE, CS, RFH, JMC, SKH

Investigation: RES, AA, ER, SL, KEM, ASB, SM, PS, MB, RP, PL, HNRS, SKH

Data Curation: RES, AA, ER, SL, KEM, RP, SKH

Visualization: RES, AA, KEM, KR

Funding acquisition: SKH

Project administration: RES, AA, ER, SL, KEM, SKH

Supervision: SKH

Writing – original draft: RES, AA, KEM, ASB, SM, PS, SKH

Writing – review & editing: RES, AA, ER, SL, KEM, KML, EBM, ASB, SM, PS, AD, CZ, VIB, SN, QC, EE, KR, MB, RP, PL, HNRS, JPD, CLS, CS, RFH, JMC, SKH

### Prior Presentation of the Information in the Manuscript

Parts of this data were presented in an oral presentation at IDWeek 2025 in October 2025 as part of the Program Committee Choice Award and as a poster at the Interdisciplinary Meeting on Antimicrobial Resistance and Innovation in January 2026. It is also available in preprint under the following citation and will later be posted to medRxiv after submission: Strength, Rachel E. and Angelova, Angelina and Robbins, Emily and PDF, Coming Into Resistance: A Longitudinal Cohort Study of Clinically Important Resistance Genes in NICU Infants Compared to Healthy Infants. Available at SSRN: https://ssrn.com/abstract=6728985 or http://dx.doi.org/10.2139/ssrn.6728985.

### Declaration of generative AI and AI-assisted technologies in the manuscript preparation process

During the preparation of this work the authors used Claude for Health and Human Services (Claude 4.5-Sonnet) and Perplexity for Government (powered by GPT-5.1) to provide suggestions to improve clarity of previously written manuscript content and find other research papers relevant to this topic. After using these tools/services, the authors reviewed and edited the content as needed and take full responsibility for the content of the published article.

