## Supplementary material for "Coming Into Resistance: Clinically Important Antibiotic Resistance Genes in NICU vs. Healthy Infants": Online-Only Supplement

### Online-Only Supplements

#### eMethods

##### +NICU Cohorts

Neonates at the Inova Fairfax Neonatal Intensive Care Unit (NICU) in Fairfax, VA were enrolled in an observational longitudinal microbiome cohort study, “Neonatal Intestinal Microbiome: Impact on Infant and Early Childhood Health and Disease,” as previously described elsewhere.[1] The study was approved by the Institutional Review Board (WCG IRB 1300205), and parental informed consent was obtained prior to enrollment. Potential participants were identified from patients in the first week of life requiring NICU admission and meeting inclusion criteria. The inclusion criteria for the infant were admission to the NICU, anticipated NICU length of stay (LOS) of 5 days or more, and maternal age of 18 years or older. Infants were excluded if they did not meet these criteria. In the NICU, stool samples were collected up to twice weekly. Detailed demographic and clinical data were collected by the study team at baseline and at the time of sampling including delivery mode, gestational age at delivery, maternal antibiotic use in pregnancy (more than 48 hours before delivery) and intrapartum (within the 48 hours before delivery), and infant antibiotic use at any time point. For this study, antibiotics refer only to those which were given orally or intravenously with systemic absorption.

After NICU discharge, parents collected stool samples every 3-6 months through approximately 36 months of corrected age using previously defined procedures.[2] Stool samples were accompanied by surveys, collecting data including health outcomes, medications, diet, and exposures. All stool samples were frozen at -80ºC until analysis.

##### -NICU Cohorts

Mothers and infants were enrolled in an IRB-approved, longitudinal cohort study "The First 1000 Days of Life and Beyond" within the Inova Health System, as previously described.[3] All experimental protocols were approved by the Inova Health System and WCG Institutional Review Board (Inova protocol #15–1804, WCG protocol #20120204), and parental informed consent was obtained prior to enrollment. The only inclusion criteria were that the parents had to be in good general health and 18 years or older; patients not meeting these criteria were excluded. Serial stool samples were collected from infants at one to two days after birth (meconium) in the well-baby nursery and at approximately two, six, twelve, and twenty-four months of age (and older, if desired). These infants were not admitted to the NICU after birth. All samples were collected using previously validated methods.[4] Demographics and clinical information were collected via questionnaires and electronic medical records review, similar to the +NICU cohorts.

##### Sample Selection

For this study, infants were included if they had at least 1 stool sample available from before 1 month of life and at least 1 stool sample available from over 1 year of age (with some exceptions, see Sample processing challenges section). Serial stool samples from the same infant had to be separated in time by more than 7 days. If more than one sample was available from a similar time period, one of the samples was chosen at random. As a result of longer hospitalization and differences in protocols, the infants with NICU exposure had more samples. Differences in sampling frequency were accounted for in later analysis.

##### DNA extraction and shotgun metagenomic sequencing

DNA was extracted from stool and sequenced using shotgun metagenomic sequencing on the NovaSeq platform as previously described.[3,5] Investigation for batch effects was performed by examining differences in read depth of all ARG (TPM values) between the different cohorts (eFigure 3). Linear mixed effect models were applied across the gene TPM values from the different cohorts, and the model was adjusted for previously discussed potential confounders. No significant batch effects were found across the ranges of TPM values of ARG in the different cohorts ($p = 0.59$). PCoA plots of beta diversity also showed overlapping distributions of NICU and non-NICU samples, demonstrating no batch-driven (i.e. cohort-related) separation (Figures 2a, 2b).

**Sample processing challenges**

33 +NICU samples were inadvertently destroyed when hardware failure occurred in the sequencing core. No patient's samples were fully lost. However, this did result in a few patients losing their post-NICU samples. Since we wanted to include as many samples in the study as possible, these participants without post-NICU samples were kept in the study. Eight +NICU samples failed sequencing thereafter. For the -NICU cohorts, due to the limited presence of bacteria around the time of birth, only 2/51 meconium samples from the -NICU cohorts had adequate sequencing data for AMR detection at the zero months of corrected age timepoint. This is not unusual for low biomass samples like meconium.[6] The two successfully sequenced meconium samples were included in the study. Missing samples and uneven distribution of samples were addressed using linear mixed models.

**Cohort Design**

To study the CI-ARG of NICU infants compared to healthy infants, we divided children from the +NICU and -NICU studies into four cohorts based on antibiotic (abx) and NICU exposure, as stated in the manuscript. To compare the groups more fairly given the long +NICU length of stay, we decided to evaluate whether infants in each of the cohorts had received antibiotics up to 2 months of corrected age (rather than just in the hospital) after we noted that 6 -NICU infants had received antibiotics within that period after discharge from the well-baby nursery (this was decided prior to final data analysis). This refinement resulted in fewer statistically significant findings on preliminary analysis. However, this approach was deemed more appropriate in capturing longitudinal CI-ARG differences in our study.

**Defining CI-ARG**

The list of CI-ARG was compiled a priori from multiple sources including the World Health Organization AWaRe (WHO Access, Watch, Reserve) Antibiotic Book, CDC 2019 Antibiotic Threats Report, and the expertise of a clinical microbiologist, infectious disease pharmacists, and infectious diseases physicians from multiple hospitals.[7,8] Bacterial ARG or groups of ARG were identified as clinically important if they had potential to cause infection resistant to empiric therapy in any age group, if they are discussed in any of the aforementioned sources, and/or if they are reasonably well-characterized in widely available antibiotic resistance gene databases. A two-level approach was used, examining both specific CI-ARG such as *mecA* or $bla_{CTX\text{-}M}$ as well as CI-ARG that confer resistance to antibiotic classes, building on prior work examining AMR by antibiotic class. In this regard, we aimed to capture both individual CI-ARG abundance trajectories over time as well as abundance trajectories of resistance to antibiotic classes. While recognizing that not all resistance genes for antibiotic classes are necessarily pathogenic or easily transferred, this two-level approach to analysis was felt to be the best way to capture both the breadth and depth of clinically important antibiotic resistance gene abundance. We also note that there is likely some overlap between resistance genes in certain antibiotic classes (e.g. penicillins and cephalosporins may have shared resistance genes), but we have listed these antibiotic class resistance genes separately for ease of interpretation.

**Sequence processing**

The quality of the raw sequencing data was assessed with FastQC and QC results summarized with MultiQC.[9,10] Adapter trimming, error correction, and quality trimming were performed by the Nephele WGSA2 pipeline, using default settings (min len=60, Ave Read Quality 10, 5' end trimming at 20, and 3' end trimming at 15).[11,12] Briefly, the WGSA2 workflow is as follows. The dataset was decontaminated using Kraken2 and a database constructed from the human and mouse genomes.[13] Taxonomic annotation and abundance estimation of the cleaned, error-corrected, and decontaminated reads were also produced with Kraken2 against the RefSeq database (March 2023).[14,15] Assembly of each sample was performed with metaSPAdes and gene prediction produced by Prodigal.[16,17] Read abundances were enumerated per scaffold by mapping cleaned, error-corrected, decontaminated reads to scaffolds. Reads mapping to gene regions were extracted from the binary alignment map (BAM) file based on the coordinates of the predicted genes and VERSE.[18] Transcripts per million (TPM) were calculated for each gene based on read counts and length of genes. After running the pipeline, the Comprehensive Antibiotic Resistance Database Resistance Gene Identifier (CARD-RGI) v.3.2.9 was used to identify which of the predicted genes were ARG.[19] Taxonomic annotations of the ARG were produced using ganon against

RefSeq (March 2023) and GTDB (r220) as a secondary database.[20,21] Taxonomic annotations of the cleaned, corrected, and decontaminated reads were also produced with ganon against RefSeq (March 2023) and the Gene Taxonomy Database (GTDB, r220) to obtain the microbial profiles of the samples. The microbial annotations from ganon were validated against the Kraken2 annotations from WGSA2 (as previously described) and the deeper taxonomic assignment used to represent the taxonomy of the gene (primarily ganon outputs).

**Corrected Age and Time Binning**

Sample collection frequency was different between the +NICU and -NICU groups as noted in the +NICU Cohorts and -NICU Cohorts sections of this supplement. Three separate methods were employed to normalize the disparities in age and sample collection frequencies between cohorts:

1. The longitudinal variable of the dataset was presented in terms of corrected age to account for both day of life (DOL, i.e. chronological age) and gestational age at birth with correction for expected full-term gestation (considered to be 40 weeks in this study). The formula to calculate corrected age was: Corrected age in weeks = (DOL/7 days/week) – (40 weeks – Gestational age at birth). Corrected-age-based models were checked and found to be similar to models using DOL plus adjustment for gestational age. Using corrected age also allowed longitudinal infant comparison with full-term, 40-week gestation as the reference point (i.e. corrected age = 0). Infants born prematurely thus had negative corrected ages early in life.
2. An in-silico, age-based binning technique was applied to the samples for some qualitative assessments (e.g. eFigures 4a, 5a, 6a). eTable 1 shows the age cutoff for the bins, and eTable 2 shows the number of samples in each bin per cohort. The age cutoffs of each bin during the premature period were decided based on prematurity intervals that roughly correspond to classification of premature infant ages per WHO guidelines.[22] For example, extremely premature infants are defined as those <28 weeks gestation, which would be -12 weeks of corrected age or younger in this study.[22] Corrected age bins after 40 weeks gestation were set around the sample collection timepoints of the -NICU infants such that the majority of -NICU infants had at least 1 sample for each age bin beyond 0 months of corrected age.
3. Samples beyond 39 months of corrected age in the +NICU groups (n=42) were excluded from the study as more diverse and complex external factors begin to affect microbiome variation later in life (e.g. social, environmental, dietary factors), and there were no samples in the -NICU groups beyond this age. This age cutoff was chosen to maximize sample retention among all groups while ensuring representation of the -NICU groups.

**ARG processing**

For all resistome exploratory methods, the ARG annotations produced by RGI were used directly, and TPM values were used for their abundance. Genes with antimicrobial resistance ontology (ARO) of interest (CI-ARGs) were identified based on the common string within gene names, families, clusters or operons, as well as drug class or antibiotic names (eTable 3). The abundance values of all genes with the same ontology of interest in an individual sample were summed to represent the abundance of CI-ARG for that sample.

**Analysis Methods Demographics**

Categorical variables among the four cohorts were compared in R version 4.5.1 using the chi-square test; chi-square tests with Monte Carlo simulation were used for comparisons where any portion of the data was not sufficiently large. Kruskal-Wallis testing was used to compare continuous variables such as gestational age and hospital length of stay.

**Resistome**

All statistical analyses were conducted and figures were produced in R version 4.5.1 with the help of RStudio, data management and visualization packages tidyverse and ggplots, specialized biological data analysis packages vegan, ampvis2, and phyloseq, and statistical packages lmerTest, rstatix, car, EnhancedVolcano, and ggsignif.

**Confounders assessment and handling**

Multiple variables were considered and tested for confounding effects on the microbiota, resistome, and CI-ARG analyses. These included demographic variables (self-identified maternal race and ethnicity, physician-assigned infant sex), maternal variables (parity, delivery mode, intrapartum antibiotics, antibiotics during pregnancy), and infant-related variables (antibiotics since last sample after two months of corrected age). Some variables (e.g. breastmilk intake) could not be formally tested or adjusted for due to insufficient data. Linear models were used to assess resistome variation and adjusted for subject identification number (SubjectID). Resistome variation was measured with the top 5 principal coordinates of the Bray-Curtis distance matrix [e.g. lmer(PCo1 ~ Cohort*Corrected_age + testConfounders + (1|SubjectID)]. Confounders were identified if the tested variable displayed more than 10% influence on the resistome response to cohorts over time. Of the tested variables, none showed any influence on resistome variation based on cohort over time; however, some showed effect on the cohort alone. To maintain consistency across resistome and microbiota analyses, all previously listed potential confounder variables were included as covariates in downstream statistical models to account for their minor but cumulative effects. Length of stay was not adjusted for since it lies on the causal pathway between NICU admission and ARG abundance. Admission to the NICU often inherently implies longer hospitalization, younger gestational age at birth, and higher levels of care than the well-baby nursery.

**Generalizability**

Recognizing that microbiomes often differ by geography and lifestyle, we aimed to recruit a relatively diverse group of participants with broad inclusion criteria to improve generalizability. Multicenter studies in a variety of locations would improve generalizability in the future.

**Diversity assessments and statistical analyses**

Richness and Shannon indices were used to assess the alpha diversity of the resistome and microbiota profiles. Gene TPM values were used as abundance measures for the resistome. Ganon-derived read counts for each taxonomy were used for microbiota profiling. Beta diversity was assessed using PCoA based on Bray-Curtis dissimilarity matrices. Linear mixed models were then used to assess microbial, resistome, and CI-ARG variation in each cohort over time. Linear mixed effects modeling was also used to assess CI-ARG based on the sums of a specified CI-ARG's abundance within each sample as described above, as well as the significant taxa representing each CI-ARG. The lines on each graph are the linear approximations of the resistance gene abundance point distributions for each cohort over time.

#### eMethods References

6. Dos Santos SJ, Pakzad Z, Elwood CN, et al. Early Neonatal Meconium Does Not Have a Demonstrable Microbiota Determined through Use of Robust Negative Controls with cpn60-Based Microbiome Profiling. *Microbiol Spectr*. 9(2):e00067-21. doi:10.1128/Spectrum.00067-21

7. *The WHO AWaRe (Access, Watch, Reserve) Antibiotic Book*. World Health Organization; 2022. Accessed December 16, 2025. https://iris.who.int/handle/10665/365237

8. CDC. 2019 Antibiotic Resistance Threats Report. Antimicrobial Resistance. February 5, 2025. Accessed March 31, 2025. https://www.cdc.gov/antimicrobial-resistance/data-research/threats/index.html

9. Andrews S. FastQC: A Quality Control Tool for High Throughput Sequence Data. https://www.bioinformatics.babraham.ac.uk/projects/fastqc/

22. *WHO Recommendations for Care of the Preterm or Low Birth Weight Infant*. 1st ed. World Health Organization; 2022.

#### eTables

##### eTable 1: Age bin minima and maxima

The minimum and maximum age for each age bin with the total number of samples in each bin.
Abbreviations: wk = weeks, m = months

| Age Bin | Total Number of Samples | Minimum Corrected Age in Bin | Maximum Corrected Age in Bin |
|---|---|---|---|
| **-3m** | 19 | -15 wk | ≤-11.5 wk |
| **-2m** | 89 | >-11.5 wk | ≤-7 wk |
| **-1m** | 197 | >-7 wk | ≤-3 wk |
| **0m** | 77 | >-3 wk | ≤1.5 wk |
| **2m** | 134 | >1.5 wk | ≤4.5 m |
| **6m** | 163 | >4.5 m | ≤11 m |
| **12m** | 185 | >11 m | ≤23 m |
| **24m** | 175 | >23 m | ≤39 m |
| **Excluded** | 42 | >39 m | |

#### eTable 2: Age bin sample distribution

This shows the number of samples per age bin per cohort.

| | Number of Samples per Cohort | | | |
|---|---|---|---|---|
| **Age Bin** | **-NICU/ -abx** | **-NICU/+abx** | **+NICU/-abx** | **+NICU/+abx** |
| **-3m** | 0 | 0 | 0 | 19 |
| **-2m** | 0 | 0 | 16 | 73 |
| **-1m** | 0 | 0 | 96 | 101 |
| **0m** | 2 | 0 | 23 | 52 |
| **2m** | 42 | 6 | 14 | 72 |
| **6m** | 48 | 6 | 54 | 55 |
| **12m** | 43 | 6 | 65 | 71 |
| **24m** | 44 | 6 | 62 | 63 |

#### eTable 3: List of CI-ARG- Gene ontologies of interest

This table lists the name of the ARG, family, or gene cluster that was examined, the antibiotic resistance ontology (ARO) that describes resistance genes or groups of genes in the Comprehensive Antibiotic Resistance Database (CARD), the number of CARD entries associated with that ARO term, the representative ontology term used for that ARO, and the number of genes found in the dataset that are associated with that ARO. Note that no mecC, VIM, KPC, IMP, or NDM beta-lactamases were identified in the samples from this study. Significantly different CI-ARG among cohorts are marked in red. The column Number of genes in CARD associated with ARO notes the values found by enumeration of the term aro_index.tsv in CARD.

| AMR Gene, Gene Family, or Gene Cluster | ARO | Number of Genes in CARD Associated with ARO | Representative Ontology Term | Number of Genes in Dataset |
|---|---|---|---|---|
| **mecA gene** | 3000617 | 1 | mecA | 1 |
| mecC gene | 3001209 | 1 | mecC | 0 |
| ampC beta-lactamase | 3000076 | 8 | ampC | 1 |
| VIM beta-lactamase | 3000021 | 72 | VIM | 0 |
| OXA-48-like beta-lactamase | 3007721 | 52 | OXA | 22 |
| IMP beta-lactamase | 3000020 | 89 | IMP | 0 |
| KPC beta-lactamase | 3000059 | 88 | KPC | 0 |
| NDM beta-lactamase | 3000057 | 47 | NDM | 0 |
| vanA gene cluster | 3000010 | 7 | VanA | 3 |
| **vanB gene cluster** | 3000013 | 8 | VanB | 7 |
| vanC gene cluster | 3000368 | 5 | VanC | 4 |
| CTX-M beta-lactamase | 3000016 | 234 | CTX-M | 6 |
| TEM beta-lactamase | 3000014 | 205 | TEM | 10 |
| SHV beta-lactamase | 3000015 | 203 | SHV | 19 |

| Antibiotic or Drug Class | ARO | Number of Genes in CARD Associated with ARO | Representative Ontology Term | Number of Genes in Dataset |
|---|---|---|---|---|
| **Aminoglycosides** | 0000016 | 309 | Aminoglycoside | 85 |
| **Fluoroquinolones** | 0000001 | 306 | Fluoroquinolone | 128 |
| **Cephalosporins** | 0000032 | 2401 | Cephalosporin | 210 |
| **Carbapenems** | 0000020 | 2340 | Carbapenem | 115 |
| **Penicillins** | 3000008 | 14 | Penicillin | 4 |
| **Macrolides** | 0000000 | 223 | Macrolide | 86 |
| Lincosamides | 0000017 | 139 | Lincosamide | 38 |
| Nitroimidazoles | 3004115 | 18 | Nitroimidazole | 12 |
| **Oxazolidinones** | 3000079 | 18 | Oxazolidinone | 6 |
| **Sulfonamides** | 3000282 | 20 | Sulfonamide | 11 |
| **Tetracyclines** | 0000051 | 244 | Tetracycline | 117 |
| Monobactams | 0000004 | 950 | Monobactam | 65 |

#### eTable 4: Post-hoc analysis of resistome alpha diversity

Exploratory, post-hoc, pairwise comparisons of resistome alpha diversity in Figure 1a between each cohort. Compared groups are shown in the two left columns; nominal p-values are shown in the rightmost column.

| Resistome Comparison | | p-value |
|---|---|---|
| -NICU/-abx | -NICU/+abx | 0.95 |
| -NICU/-abx | +NICU/-abx | <0.001 |
| -NICU/-abx | +NICU/+abx | <0.001 |
| -NICU/+abx | +NICU/-abx | <0.001 |
| -NICU/+abx | +NICU/+abx | <0.001 |
| +NICU/-abx | +NICU/+abx | 0.56 |

#### eTable 5: Post-hoc analysis of resistome beta diversity

Exploratory, post-hoc, pairwise comparisons of resistome beta diversity (PCo1) in Figure 2a between each cohort. Compared groups are shown in the two left columns; effect sizes and nominal p-values are shown in the two right columns.

| Resistome Comparison | | PCo1 Effect Size | PCo1 p-value |
|---|---|---|---|
| -NICU/-abx | -NICU/+abx | 0.0097 | 0.70 |
| -NICU/-abx | +NICU/-abx | 0.34 | <0.001 |
| -NICU/-abx | +NICU/+abx | 0.23 | <0.001 |
| -NICU/+abx | +NICU/-abx | 0.20 | 0.001 |
| -NICU/+abx | +NICU/+abx | 0.21 | <0.001 |
| +NICU/-abx | +NICU/+abx | 0.0002 | 0.69 |

#### eTable 6: Post-hoc comparison of CI-ARG by NICU exposure

Exploratory, post-hoc comparisons of CI-ARG in Figure 3 by NICU exposure are shown here. Effect sizes and nominal p-values are shown for comparisons between infants with and without NICU exposure for each antibiotic shown in Figure 3, pooling across early-life antibiotic exposure status. For instance, cephalosporin trajectories differed significantly by NICU exposure (p<0.001).

| CI-ARG Comparison, +NICU vs –NICU | Effect Size | p-value |
|---|---|---|
| Cephalosporin | 0.46 | <0.001 |
| Oxazolidinone | 0.46 | <0.001 |
| Sulfonamide | 0.42 | 0.16 |
| *vanB* | 0.43 | <0.001 |
| Aminoglycoside | 0.42 | <0.001 |
| Penicillin | 0.41 | 0.007 |
| Macrolide | 0.41 | <0.001 |
| Carbapenem | 0.35 | 0.004 |
| Fluoroquinolone | 0.35 | <0.001 |
| *mecA* | 0.25 | <0.001 |
| Tetracycline | 0.20 | <0.001 |

#### eTable 7: Post-hoc comparison of CI-ARG by early-life antibiotic exposure

Exploratory, post-hoc comparisons of CI-ARG in Figure 3 by antibiotic exposure are shown here. Effect sizes and nominal p-values are shown for comparisons between infants with and without early-life antibiotic exposure for each antibiotic shown in Figure 3, pooling across NICU status. For instance, cephalosporin trajectories for infants with early-life antibiotic exposure do not differ significantly from infants without antibiotic exposure (p=0.66).

| CI-ARG Comparison, +abx vs -abx | Effect Size | p-value |
|---|---|---|
| Cephalosporin | 0.45 | 0.66 |
| Oxazolidinone | 0.42 | 0.076 |
| Sulfonamide | 0.43 | 0.005 |
| *vanB* | 0.41 | 0.51 |
| Aminoglycoside | 0.40 | 0.27 |
| Penicillin | 0.41 | 0.75 |
| Macrolide | 0.39 | 0.51 |
| Carbapenem | 0.35 | 0.51 |
| Fluoroquinolone | 0.33 | 0.57 |
| *mecA* | 0.23 | 0.62 |
| Tetracycline | 0.18 | 0.78 |

### eFigures Captions

#### eFigures 1a-1b: Principal coordinates analysis of Bray-Curtis beta diversity by age bin

(a) Overall resistome. Each point represents a sample's resistome within a specific age bin, denoted by the different colors. The nominal p-values compare the resistomes of the cohorts to each other within one age bin and are corrected for tested confounders. For instance, the two-month samples of the cohorts are significantly different from each other (p=0.001). The abbreviation NS means not significant. The ellipses contain the centroids of all samples in each age bin. Major ARG contributors to the distribution of the dots on the ordination space are shown in the rectangles on the graph. Note that major ARG contributors are different in color and do not correspond to any particular age bin. (b) Overall Microbiota. Each point represents a sample's microbiota within a specific age bin, denoted by color. The nominal p-values compare the microbiota of the cohorts to each other within one age bin and are corrected for tested confounders. The ellipses again contain the centroids of all samples in each age bin. Major taxonomic contributors per age bin are listed in the rectangles where applicable; colors do not correspond to any particular age bin.

#### eFigure 2: Most abundant ARG across age bins per cohort

This figure shows the most abundant ARG in the dataset regardless of clinical importance, along with the antibiotic classes with which they are associated. Abundance in TPM is shown by both number and color.

#### eFigure 3: Sequencing depths across cohorts

This box plot denotes the abundance values in transcripts per million (TPM) of all ARG in each cohort, indicating that the cohorts did not have a significantly different range of TPM values representing the ARG (p=0.59), suggesting no batch effect. The accompanying table assesses the significance of the cohort variable adjusted for all other variables and is shown for completeness. The Chisq column shows the chi-square statistic, Df shows degrees of freedom, and Sig shows which values are significant.

#### eFigures 4a-4d: vanB analysis over time between cohorts

(4a) Participant-level *vanB* gene abundance over time. Each cohort is listed on the left side of the plots, and each line within the graphs represents one participant. The participant's samples are denoted by the diamonds plotted on those lines. Age bins are listed above the graphs with age bin limits marked with thin, vertical lines across all graphs (age bin limits in eTable 1). The vertical, blue, dotted line marks 0 weeks of corrected age. The abundance of the resistance gene in that sample is shown by color, with more yellow coloring indicating higher abundance. Horizontal gray bars represent the duration of hospitalization, and the vertical gray line at the end of each horizontal bar represents the time of discharge for that participant. The horizontal red lines indicate times where antibiotics were given in the NICU; post-discharge from the hospital, the red lines indicate that antibiotics were given prior to that sample. Gray diamonds indicate that the resistance gene of interest was not detected in that sample. (4b) Top species carrying *vanB*. Species carrying significantly different amounts of the resistance gene across cohorts are denoted by asterisks. Age bins in gray did not have samples for the listed cohort. Abundance of the resistance gene in transcripts per million is shown by the color of the rectangles for each taxon. This was calculated with linear mixed effects models for each taxon and adjusted for possible confounders. (4c) Longitudinal abundance of species carrying significantly different amounts of *vanB* by cohort. This was calculated with linear mixed effects differential abundance analysis and corrected for all tested confounders. (4d) Abundance of each taxon from eFigure 4c regardless of *vanB* carriage.

#### eFigures 5a-5d: Aminoglycoside resistance gene analysis over time between cohorts.

(5a) Participant-level aminoglycoside ARG abundance over time. See eFigure 4a caption for reference. (5b) Top species carrying aminoglycoside ARG. See eFigure 4b caption for reference. (5c) Longitudinal abundance of species carrying significantly different amounts of aminoglycoside ARG by cohort. (5d) Abundance of each taxon from eFigure 5c regardless of aminoglycoside ARG carriage.

##### eFigures 6a-6d: mecA analysis over time between cohorts

(6a) Participant-level *mecA* abundance over time. See eFigure 4a caption for reference. (6b) Top species carrying *mecA*. See eFigure 4b caption for reference. (6c) Longitudinal abundance of species carrying significantly different amounts of *mecA* by cohort. (6d) Abundance of each taxon from eFigure 6c regardless of *mecA* carriage.

##### eFigure 7: Participant-level oxazolidinone resistance gene abundance over time

Oxazolidinone resistance gene abundance was significantly different among the four cohorts. See eFigure 4a caption for reference.

##### eFigure 8: Participant-level penicillin resistance gene abundance over time

Penicillin resistance gene abundance was significantly different among the four cohorts. See eFigure 4a caption for reference.

##### eFigure 9: Participant-level cephalosporin resistance gene abundance over time

Cephalosporin resistance gene abundance was significantly different among the four cohorts. See eFigure 4a caption for reference.

##### eFigure 10: Participant-level macrolide resistance gene abundance over time

Macrolide resistance gene abundance was significantly different among the four cohorts. See eFigure 4a caption for reference.

##### eFigure 11: Participant-level carbapenem resistance gene abundance over time

Carbapenem resistance gene abundance was significantly different among the four cohorts. See eFigure 4a caption for reference.

##### eFigure 12: Participant-level fluoroquinolone resistance gene abundance over time

Fluoroquinolone resistance gene abundance was significantly different among the four cohorts. See eFigure 4a caption for reference.

##### eFigure 13: Participant-level tetracycline resistance gene abundance over time

Tetracycline resistance gene abundance was significantly different among the four cohorts. See eFigure 4a caption for reference.

##### eFigure 14: Participant-level sulfonamide resistance gene abundance over time

Sulfonamide resistance gene abundance was significantly different among the four cohorts. See eFigure 4a caption for reference.

##### eFigure 15: Participant-level $bla_{ampC}$ abundance over time

$bla_{ampC}$ abundance was not significantly different among the four cohorts. See eFigure 4a caption for reference.

##### eFigure 16: Participant-level $bla_{OXA-48}$-like gene abundance over time

$bla_{OXA-48}$-like gene abundance was not significantly different among the four cohorts. See eFigure 4a caption for reference.

##### eFigure 17: Participant-level *vanA* abundance over time

*vanA* abundance was not significantly different among the four cohorts. See eFigure 4a caption for reference.

##### eFigure 18: Participant-level *vanC* abundance over time
*vanC* abundance was not significantly different among the four cohorts. See eFigure 4a caption for reference.

##### eFigure 19: Participant-level *bla*$_{CTX\text{-}M}$ abundance over time
*bla*CTX-M abundance was not significantly different among the four cohorts. See eFigure 4a caption for reference.

##### eFigure 20: Participant-level *bla*$_{TEM}$ abundance over time
*bla*TEM abundance was not significantly different among the four cohorts. See eFigure 4a caption for reference.

##### eFigure 21: Participant-level *bla*$_{SHV}$ abundance over time
*bla*SHV abundance was not significantly different among the four cohorts. See eFigure 4a caption for reference.

##### eFigure 22: Participant-level lincosamide resistance gene abundance over time
Lincosamide resistance gene abundance was not significantly different among the four cohorts. See eFigure 4a caption for reference.

##### eFigure 23: Participant-level nitroimidazole resistance gene abundance over time
Nitroimidazole resistance gene abundance was not significantly different among the four cohorts. See eFigure 4a caption for reference.

##### eFigure 24: Participant-level monobactam resistance gene abundance over time
Monobactam resistance gene abundance was not significantly different among the four cohorts. See eFigure 4a caption for reference.

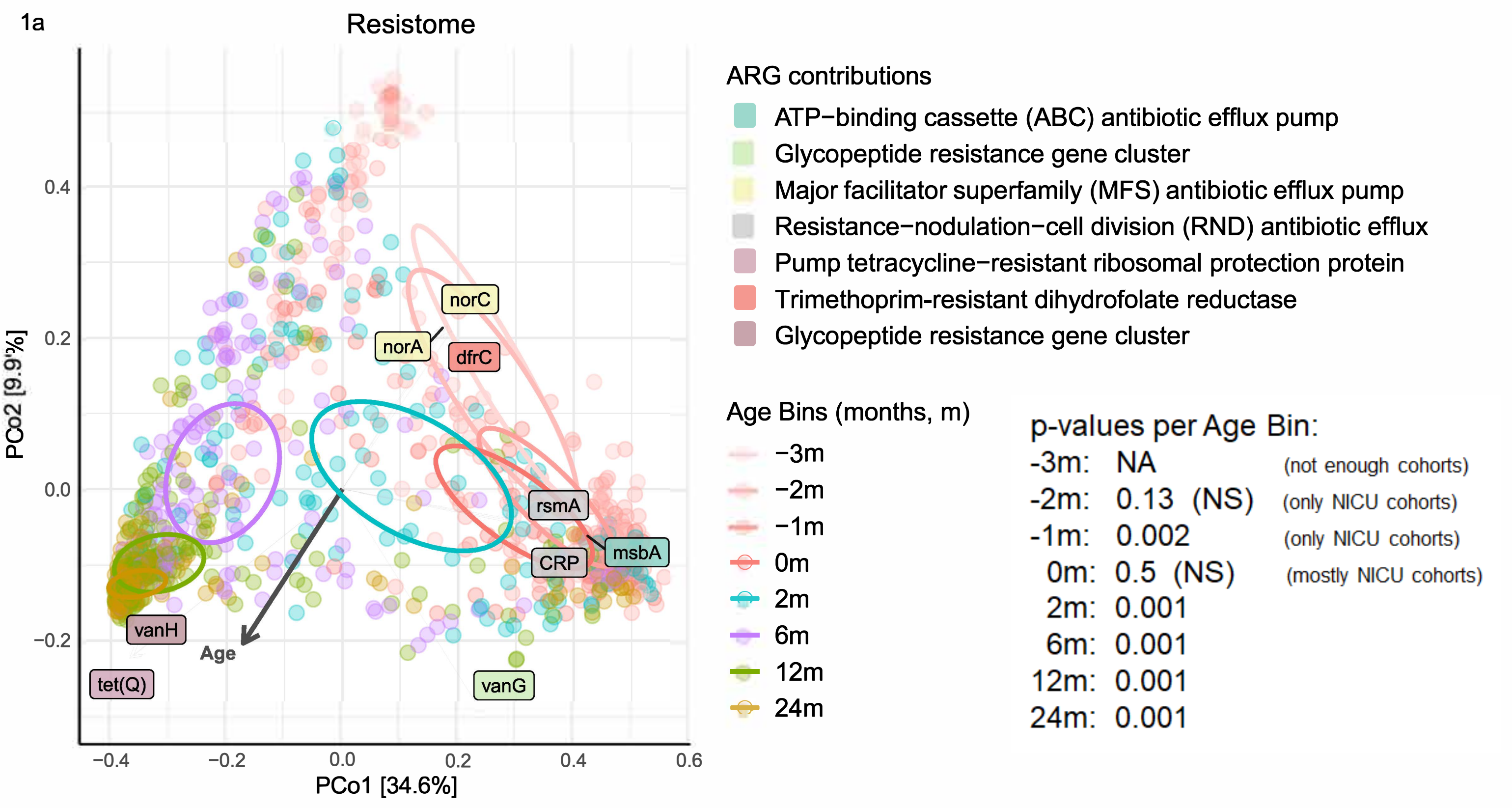
1a
Resistome
ARG contributions
ATP−binding cassette (ABC) antibiotic efflux pump
Glycopeptide resistance gene cluster
Major facilitator superfamily (MFS) antibiotic efflux pump
Resistance−nodulation−cell division (RND) antibiotic efflux
Pump tetracycline−resistant ribosomal protection protein
Trimethoprim-resistant dihydrofolate reductase
Glycopeptide resistance gene cluster
Age Bins (months, m)
−3m
−2m
−1m
0m
2m
6m
12m
24m
p-values per Age Bin:
-3m: NA (not enough cohorts)
-2m: 0.13 (NS) (only NICU cohorts)
-1m: 0.002 (only NICU cohorts)
0m: 0.5 (NS) (mostly NICU cohorts)
2m: 0.001
6m: 0.001
12m: 0.001
24m: 0.001
norC
norA
dfrC
rsmA
msbA
CRP
vanH
Age
tet(Q)
vanG
PCo2 [9.9%]
PCo1 [34.6%]
0.4
0.2
0.0
−0.2
−0.4
−0.2
0.0
0.2
0.4
0.6

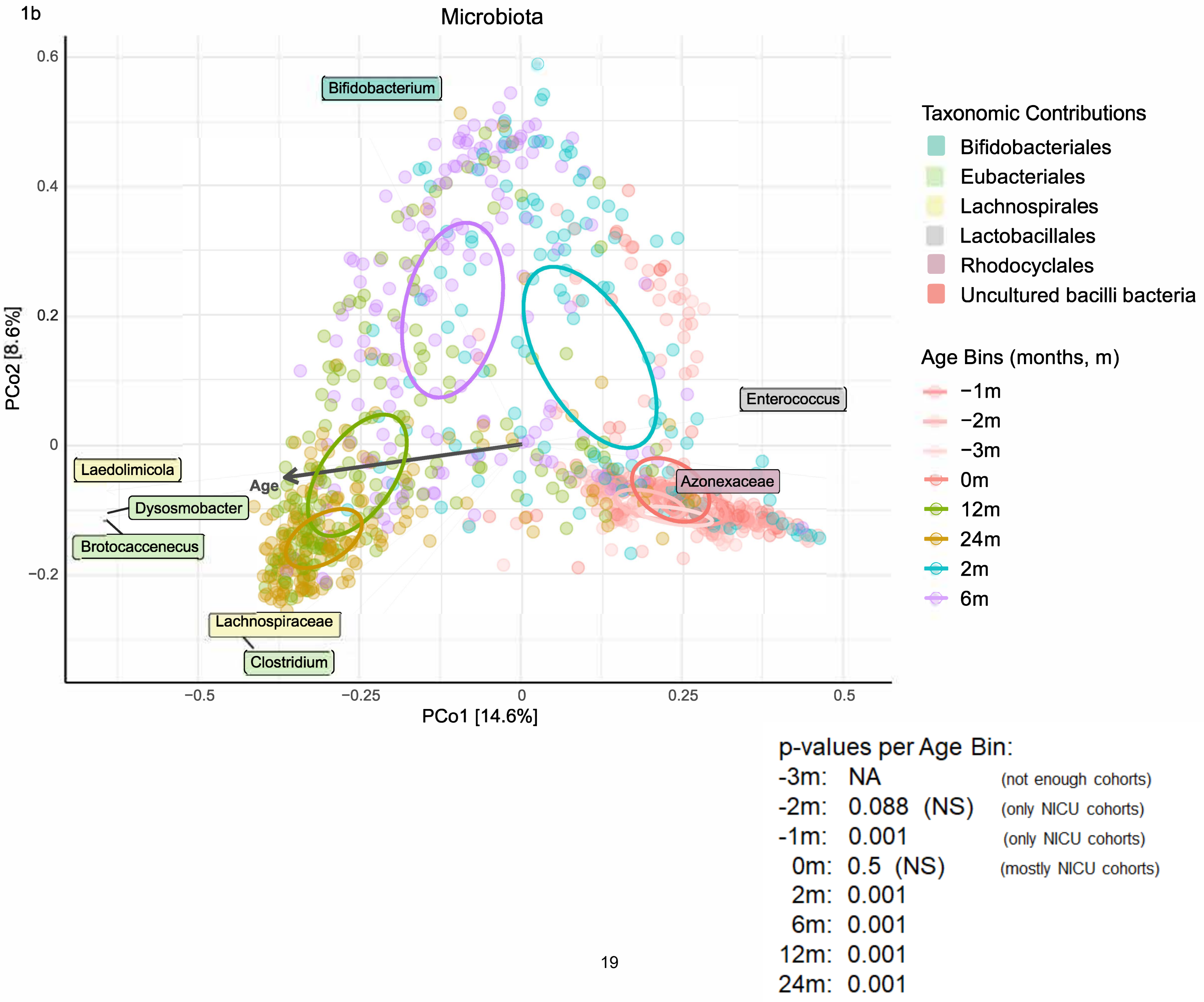
1b
Microbiota
Bifidobacterium
Enterococcus
Azonexaceae
Laedolimicola
Dysosmobacter
Brotocaccenecus
Lachnospiraceae
Clostridium
Age
Taxonomic Contributions
Bifidobacteriales
Eubacteriales
Lachnospirales
Lactobacillales
Rhodocyclales
Uncultured bacilli bacteria
Age Bins (months, m)
−1m
−2m
−3m
0m
12m
24m
2m
6m
PCo2 [8.6%]
PCo1 [14.6%]
0.6
0.4
0.2
0
−0.2
−0.5
−0.25
0
0.25
0.5
p-values per Age Bin:
-3m: NA (not enough cohorts)
-2m: 0.088 (NS) (only NICU cohorts)
-1m: 0.001 (only NICU cohorts)
0m: 0.5 (NS) (mostly NICU cohorts)
2m: 0.001
6m: 0.001
12m: 0.001
24m: 0.001

2

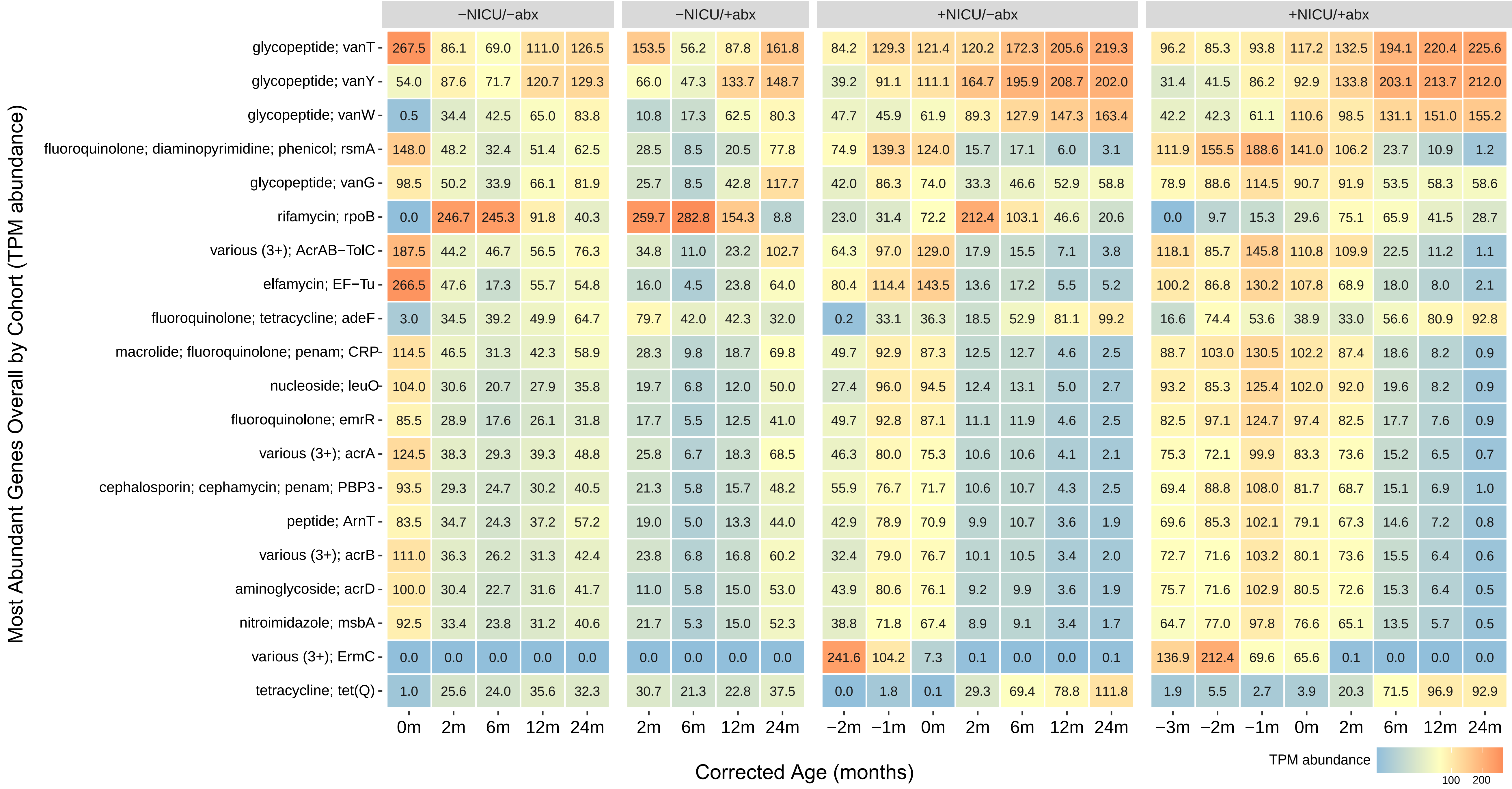

−NICU/−abx
−NICU/+abx
+NICU/−abx
+NICU/+abx
Most Abundant Genes Overall by Cohort (TPM abundance)
glycopeptide; vanT 267.5 86.1 69.0 111.0 126.5 153.5 56.2 87.8 161.8 84.2 129.3 121.4 120.2 172.3 205.6 219.3 96.2 85.3 93.8 117.2 132.5 194.1 220.4 225.6
glycopeptide; vanY 54.0 87.6 71.7 120.7 129.3 66.0 47.3 133.7 148.7 39.2 91.1 111.1 164.7 195.9 208.7 202.0 31.4 41.5 86.2 92.9 133.8 203.1 213.7 212.0
glycopeptide; vanW 0.5 34.4 42.5 65.0 83.8 10.8 17.3 62.5 80.3 47.7 45.9 61.9 89.3 127.9 147.3 163.4 42.2 42.3 61.1 110.6 98.5 131.1 151.0 155.2
fluoroquinolone; diaminopyrimidine; phenicol; rsmA 148.0 48.2 32.4 51.4 62.5 28.5 8.5 20.5 77.8 74.9 139.3 124.0 15.7 17.1 6.0 3.1 111.9 155.5 188.6 141.0 106.2 23.7 10.9 1.2
glycopeptide; vanG 98.5 50.2 33.9 66.1 81.9 25.7 8.5 42.8 117.7 42.0 86.3 74.0 33.3 46.6 52.9 58.8 78.9 88.6 114.5 90.7 91.9 53.5 58.3 58.6
rifamycin; rpoB 0.0 246.7 245.3 91.8 40.3 259.7 282.8 154.3 8.8 23.0 31.4 72.2 212.4 103.1 46.6 20.6 0.0 9.7 15.3 29.6 75.1 65.9 41.5 28.7
various (3+); AcrAB−TolC 187.5 44.2 46.7 56.5 76.3 34.8 11.0 23.2 102.7 64.3 97.0 129.0 17.9 15.5 7.1 3.8 118.1 85.7 145.8 110.8 109.9 22.5 11.2 1.1
elfamycin; EF−Tu 266.5 47.6 17.3 55.7 54.8 16.0 4.5 23.8 64.0 80.4 114.4 143.5 13.6 17.2 5.5 5.2 100.2 86.8 130.2 107.8 68.9 18.0 8.0 2.1
fluoroquinolone; tetracycline; adeF 3.0 34.5 39.2 49.9 64.7 79.7 42.0 42.3 32.0 0.2 33.1 36.3 18.5 52.9 81.1 99.2 16.6 74.4 53.6 38.9 33.0 56.6 80.9 92.8
macrolide; fluoroquinolone; penam; CRP 114.5 46.5 31.3 42.3 58.9 28.3 9.8 18.7 69.8 49.7 92.9 87.3 12.5 12.7 4.6 2.5 88.7 103.0 130.5 102.2 87.4 18.6 8.2 0.9
nucleoside; leuO 104.0 30.6 20.7 27.9 35.8 19.7 6.8 12.0 50.0 27.4 96.0 94.5 12.4 13.1 5.0 2.7 93.2 85.3 125.4 102.0 92.0 19.6 8.2 0.9
fluoroquinolone; emrR 85.5 28.9 17.6 26.1 31.8 17.7 5.5 12.5 41.0 49.7 92.8 87.1 11.1 11.9 4.6 2.5 82.5 97.1 124.7 97.4 82.5 17.7 7.6 0.9
various (3+); acrA 124.5 38.3 29.3 39.3 48.8 25.8 6.7 18.3 68.5 46.3 80.0 75.3 10.6 10.6 4.1 2.1 75.3 72.1 99.9 83.3 73.6 15.2 6.5 0.7
cephalosporin; cephamycin; penam; PBP3 93.5 29.3 24.7 30.2 40.5 21.3 5.8 15.7 48.2 55.9 76.7 71.7 10.6 10.7 4.3 2.5 69.4 88.8 108.0 81.7 68.7 15.1 6.9 1.0
peptide; ArnT 83.5 34.7 24.3 37.2 57.2 19.0 5.0 13.3 44.0 42.9 78.9 70.9 9.9 10.7 3.6 1.9 69.6 85.3 102.1 79.1 67.3 14.6 7.2 0.8
various (3+); acrB 111.0 36.3 26.2 31.3 42.4 23.8 6.8 16.8 60.2 32.4 79.0 76.7 10.1 10.5 3.4 2.0 72.7 71.6 103.2 80.1 73.6 15.5 6.4 0.6
aminoglycoside; acrD 100.0 30.4 22.7 31.6 41.7 11.0 5.8 15.0 53.0 43.9 80.6 76.1 9.2 9.9 3.6 1.9 75.7 71.6 102.9 80.5 72.6 15.3 6.4 0.5
nitroimidazole; msbA 92.5 33.4 23.8 31.2 40.6 21.7 5.3 15.0 52.3 38.8 71.8 67.4 8.9 9.1 3.4 1.7 64.7 77.0 97.8 76.6 65.1 13.5 5.7 0.5
various (3+); ErmC 0.0 0.0 0.0 0.0 0.0 0.0 0.0 0.0 0.0 241.6 104.2 7.3 0.1 0.0 0.0 0.1 136.9 212.4 69.6 65.6 0.1 0.0 0.0 0.0
tetracycline; tet(Q) 1.0 25.6 24.0 35.6 32.3 30.7 21.3 22.8 37.5 0.0 1.8 0.1 29.3 69.4 78.8 111.8 1.9 5.5 2.7 3.9 20.3 71.5 96.9 92.9
0m 2m 6m 12m 24m
2m 6m 12m 24m
−2m −1m 0m 2m 6m 12m 24m
−3m −2m −1m 0m 2m 6m 12m 24m
Corrected Age (months)
TPM abundance
100
200

3

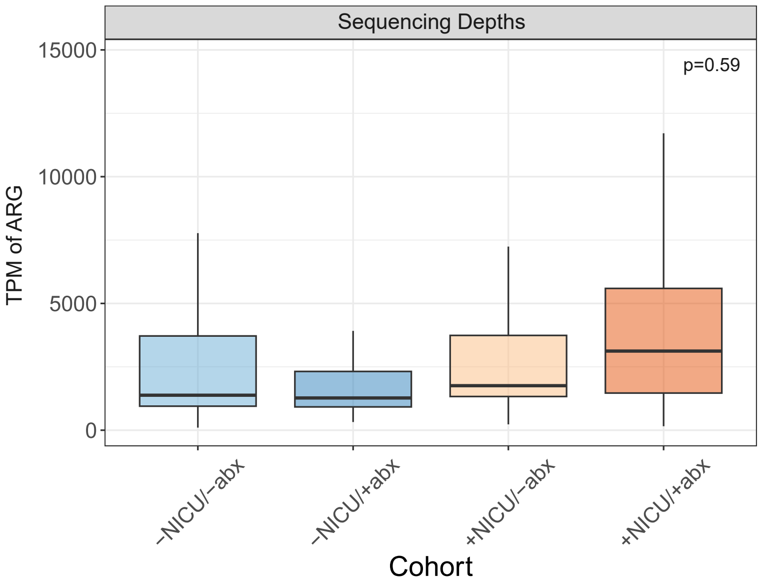

| Predictor | Chisq | Df | P-value | Sig |
|---|---|---|---|---|
| (Intercept) | 18·86 | 1 | <0·0001 | *** |
| Cohort | 1·9 | 3 | 0·59 | |
| Corrected age (weeks) | 207·31 | 1 | <0·0001 | *** |
| Parity | 0·01 | 1 | 0·91 | |
| Race | 2·32 | 3 | 0·51 | |
| Ethnicity | 0·59 | 2 | 0·74 | |
| Sex | 0·87 | 1 | 0·35 | |
| Delivery mode | 4·99 | 1 | 0·025 | * |
| Intrapartum antibiotics | 5·18 | 1 | 0·023 | * |
| Antibiotics during pregnancy | 1·14 | 1 | 0·29 | |
| Antibiotics since last sample | 44·54 | 1 | <0·0001 | *** |

4a

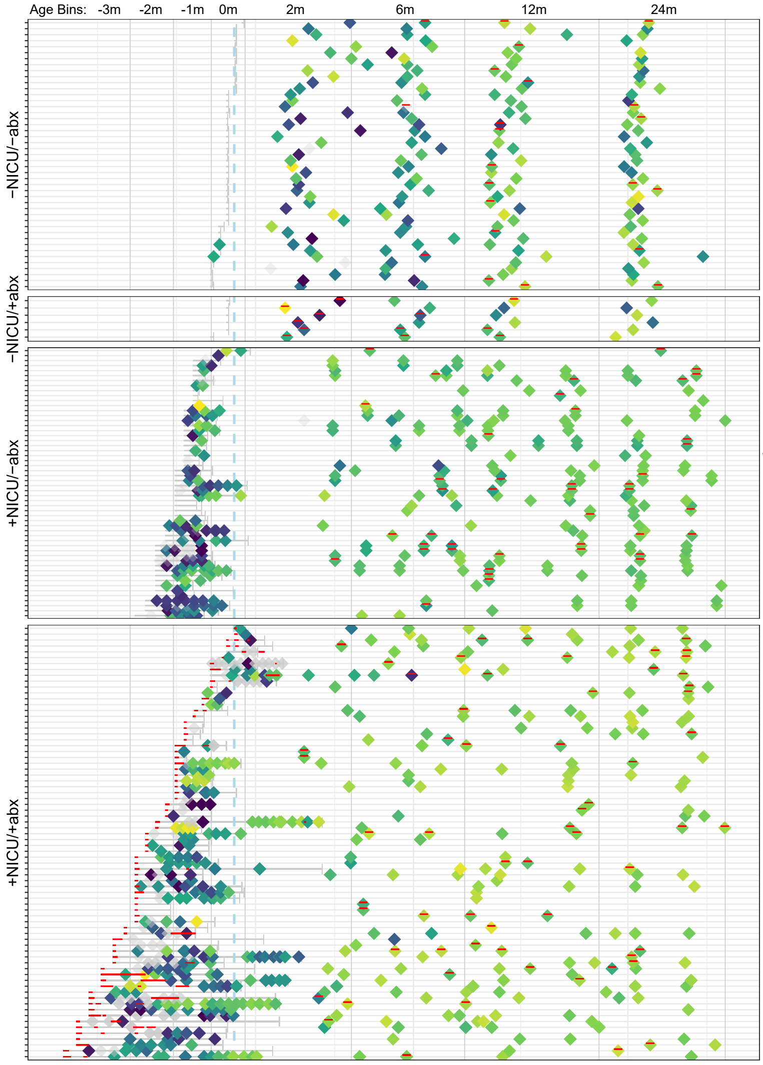
vanB
Age Bins: -3m -2m -1m 0m 2m 6m 12m 24m
−NICU/−abx
−NICU/+abx
+NICU/−abx
+NICU/+abx
Abundance (TPM)
400
300
200
100

4c

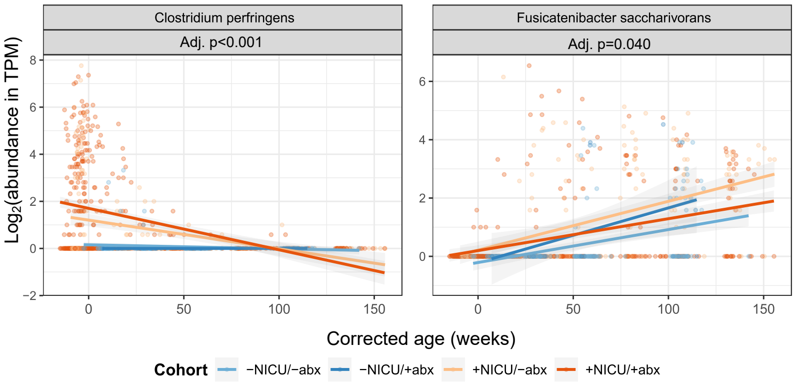
Clostridium perfringens
Adj. p<0.001
Fusicatenibacter saccharivorans
Adj. p=0.040
Log2(abundance in TPM)
Corrected age (weeks)
0 50 100 150
Cohort −NICU/−abx −NICU/+abx +NICU/−abx +NICU/+abx

4d

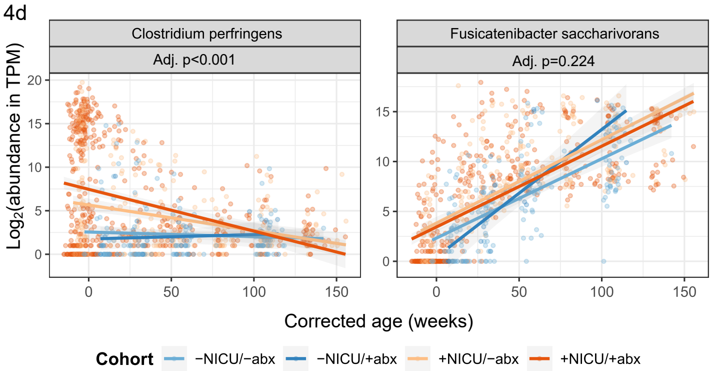
Clostridium perfringens
Adj. p<0.001
Fusicatenibacter saccharivorans
Adj. p=0.224
Log2(abundance in TPM)
Corrected age (weeks)
0 50 100 150
Cohort −NICU/−abx −NICU/+abx +NICU/−abx +NICU/+abx

4b

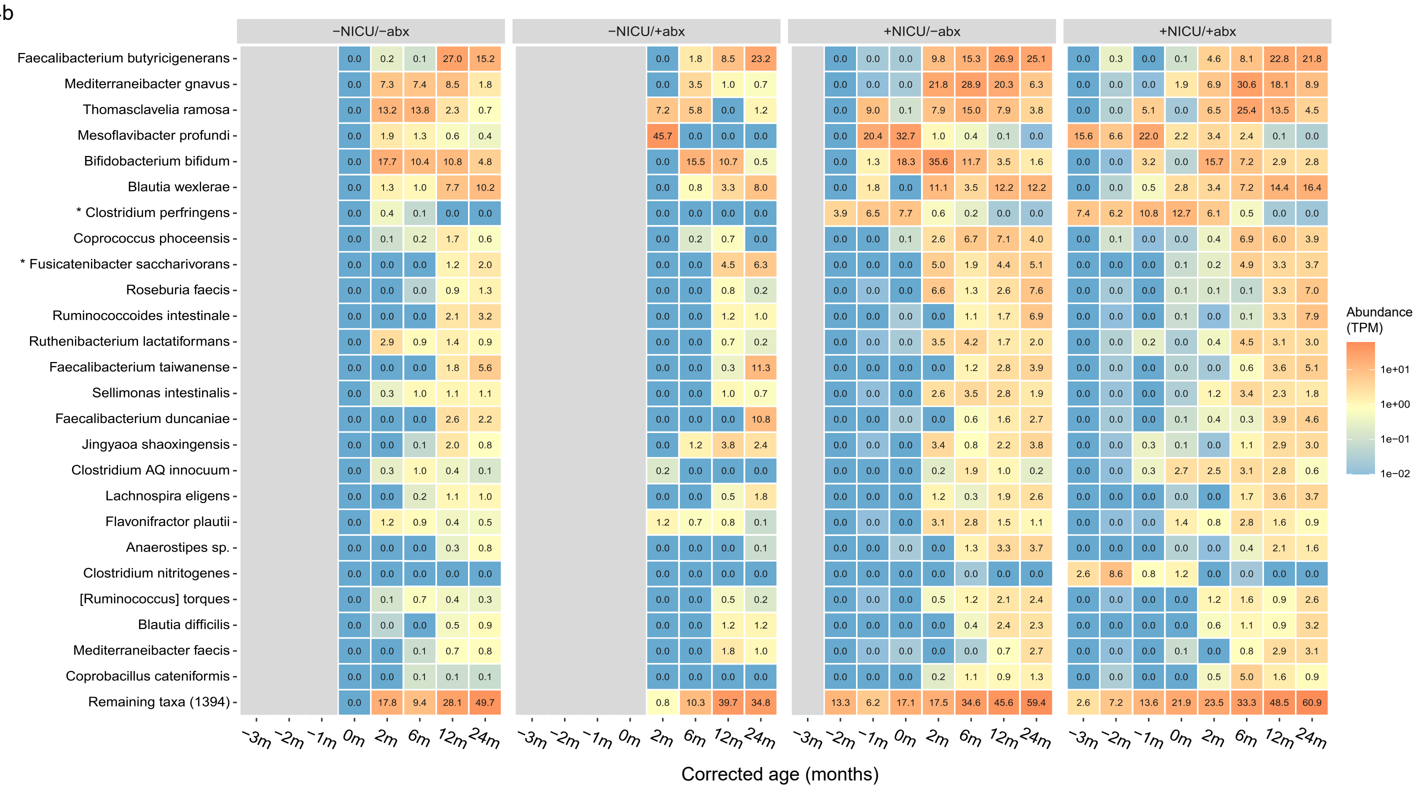
−NICU/−abx
−NICU/+abx
+NICU/−abx
+NICU/+abx
Faecalibacterium butyricigenerans - 0.0 0.2 0.1 27.0 15.2 | 0.0 1.8 8.5 23.2 | 0.0 0.0 0.0 9.8 15.3 26.9 25.1 | 0.0 0.3 0.0 0.1 4.6 8.1 22.8 21.8
Mediterraneibacter gnavus - 0.0 7.3 7.4 8.5 1.8 | 0.0 3.5 1.0 0.7 | 0.0 0.0 0.0 21.8 28.9 20.3 6.3 | 0.0 0.0 0.0 1.9 6.9 30.6 18.1 8.9
Thomasclavelia ramosa - 0.0 13.2 13.8 2.3 0.7 | 7.2 5.8 0.0 1.2 | 0.0 9.0 0.1 7.9 15.0 7.9 3.8 | 0.0 0.0 5.1 0.0 6.5 25.4 13.5 4.5
Mesoflavibacter profundi - 0.0 1.9 1.3 0.6 0.4 | 45.7 0.0 0.0 0.0 | 0.0 20.4 32.7 1.0 0.4 0.1 0.0 | 15.6 6.6 22.0 2.2 3.4 2.4 0.1 0.0
Bifidobacterium bifidum - 0.0 17.7 10.4 10.8 4.8 | 0.0 15.5 10.7 0.5 | 0.0 1.3 18.3 35.6 11.7 3.5 1.6 | 0.0 0.0 3.2 0.0 15.7 7.2 2.9 2.8
Blautia wexlerae - 0.0 1.3 1.0 7.7 10.2 | 0.0 0.8 3.3 8.0 | 0.0 1.8 0.0 11.1 3.5 12.2 12.2 | 0.0 0.0 0.5 2.8 3.4 7.2 14.4 16.4
* Clostridium perfringens - 0.0 0.4 0.1 0.0 0.0 | 0.0 0.0 0.0 0.0 | 3.9 6.5 7.7 0.6 0.2 0.0 0.0 | 7.4 6.2 10.8 12.7 6.1 0.5 0.0 0.0
Coprococcus phoceensis - 0.0 0.1 0.2 1.7 0.6 | 0.0 0.2 0.7 0.0 | 0.0 0.0 0.1 2.6 6.7 7.1 4.0 | 0.0 0.1 0.0 0.0 0.4 6.9 6.0 3.9
* Fusicatenibacter saccharivorans - 0.0 0.0 0.0 1.2 2.0 | 0.0 0.0 4.5 6.3 | 0.0 0.0 0.0 5.0 1.9 4.4 5.1 | 0.0 0.0 0.0 0.1 0.2 4.9 3.3 3.7
Roseburia faecis - 0.0 0.0 0.0 0.9 1.3 | 0.0 0.0 0.8 0.2 | 0.0 0.0 0.0 6.6 1.3 2.6 7.6 | 0.0 0.0 0.0 0.1 0.1 0.1 3.3 7.0
Ruminococcoides intestinale - 0.0 0.0 0.0 2.1 3.2 | 0.0 0.0 1.2 1.0 | 0.0 0.0 0.0 0.0 1.1 1.7 6.9 | 0.0 0.0 0.0 0.1 0.0 0.1 3.3 7.9
Ruthenibacterium lactatiformans - 0.0 2.9 0.9 1.4 0.9 | 0.0 0.0 0.7 0.2 | 0.0 0.0 0.0 3.5 4.2 1.7 2.0 | 0.0 0.0 0.2 0.0 0.4 4.5 3.1 3.0
Faecalibacterium taiwanense - 0.0 0.0 0.0 1.8 5.6 | 0.0 0.0 0.3 11.3 | 0.0 0.0 0.0 0.0 1.2 2.8 3.9 | 0.0 0.0 0.0 0.0 0.0 0.6 3.6 5.1
Sellimonas intestinalis - 0.0 0.3 1.0 1.1 1.1 | 0.0 0.0 1.0 0.7 | 0.0 0.0 0.0 2.6 3.5 2.8 1.9 | 0.0 0.0 0.0 0.0 1.2 3.4 2.3 1.8
Faecalibacterium duncaniae - 0.0 0.0 0.0 2.6 2.2 | 0.0 0.0 0.0 10.8 | 0.0 0.0 0.0 0.0 0.6 1.6 2.7 | 0.0 0.0 0.0 0.1 0.4 0.3 3.9 4.6
Jingyaoa shaoxingensis - 0.0 0.0 0.1 2.0 0.8 | 0.0 1.2 3.8 2.4 | 0.0 0.0 0.0 3.4 0.8 2.2 3.8 | 0.0 0.0 0.3 0.1 0.0 1.1 2.9 3.0
Clostridium AQ innocuum - 0.0 0.3 1.0 0.4 0.1 | 0.2 0.0 0.0 0.0 | 0.0 0.0 0.0 0.2 1.9 1.0 0.2 | 0.0 0.0 0.3 2.7 2.5 3.1 2.8 0.6
Lachnospira eligens - 0.0 0.0 0.2 1.1 1.0 | 0.0 0.0 0.5 1.8 | 0.0 0.0 0.0 1.2 0.3 1.9 2.6 | 0.0 0.0 0.0 0.0 0.0 1.7 3.6 3.7
Flavonifractor plautii - 0.0 1.2 0.9 0.4 0.5 | 1.2 0.7 0.8 0.1 | 0.0 0.0 0.0 3.1 2.8 1.5 1.1 | 0.0 0.0 0.0 1.4 0.8 2.8 1.6 0.9
Anaerostipes sp. - 0.0 0.0 0.0 0.3 0.8 | 0.0 0.0 0.0 0.1 | 0.0 0.0 0.0 0.0 1.3 3.3 3.7 | 0.0 0.0 0.0 0.0 0.0 0.4 2.1 1.6
Clostridium nitritogenes - 0.0 0.0 0.0 0.0 0.0 | 0.0 0.0 0.0 0.0 | 0.0 0.0 0.0 0.0 0.0 0.0 0.0 | 2.6 8.6 0.8 1.2 0.0 0.0 0.0 0.0
[Ruminococcus] torques - 0.0 0.1 0.7 0.4 0.3 | 0.0 0.0 0.5 0.2 | 0.0 0.0 0.0 0.5 1.2 2.1 2.4 | 0.0 0.0 0.0 0.0 1.2 1.6 0.9 2.6
Blautia difficilis - 0.0 0.0 0.0 0.5 0.9 | 0.0 0.0 1.2 1.2 | 0.0 0.0 0.0 0.0 0.4 2.4 2.3 | 0.0 0.0 0.0 0.0 0.6 1.1 0.9 3.2
Mediterraneibacter faecis - 0.0 0.0 0.1 0.7 0.8 | 0.0 0.0 1.8 1.0 | 0.0 0.0 0.0 0.0 0.0 0.7 2.7 | 0.0 0.0 0.0 0.1 0.0 0.8 2.9 3.1
Coprobacillus cateniformis - 0.0 0.0 0.1 0.1 0.1 | 0.0 0.0 0.0 0.0 | 0.0 0.0 0.0 0.2 1.1 0.9 1.3 | 0.0 0.0 0.0 0.0 0.5 5.0 1.6 0.9
Remaining taxa (1394) - 0.0 17.8 9.4 28.1 49.7 | 0.8 10.3 39.7 34.8 | 13.3 6.2 17.1 17.5 34.6 45.6 59.4 | 2.6 7.2 13.6 21.9 23.5 33.3 48.5 60.9
−3m −2m −1m 0m 2m 6m 12m 24m
Corrected age (months)
Abundance (TPM)
1e+01
1e+00
1e−01
1e−02

5a

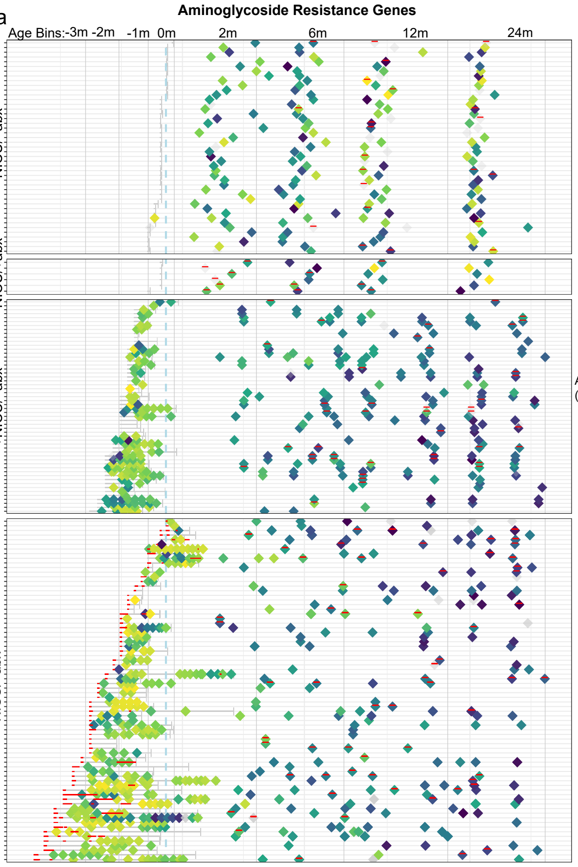
Aminoglycoside Resistance Genes
Age Bins: -3m -2m -1m 0m 2m 6m 12m 24m
−NICU/−abx
−NICU/+abx
+NICU/−abx
+NICU/+abx
Abundance (TPM)
1500
1000
500

5c

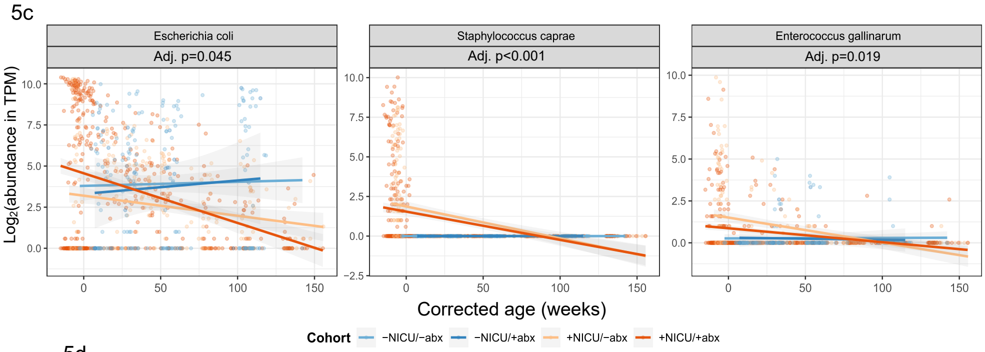
Escherichia coli
Adj. p=0.045
Staphylococcus caprae
Adj. p<0.001
Enterococcus gallinarum
Adj. p=0.019
Log2(abundance in TPM)
Corrected age (weeks)
Cohort
−NICU/−abx
−NICU/+abx
+NICU/−abx
+NICU/+abx

5d

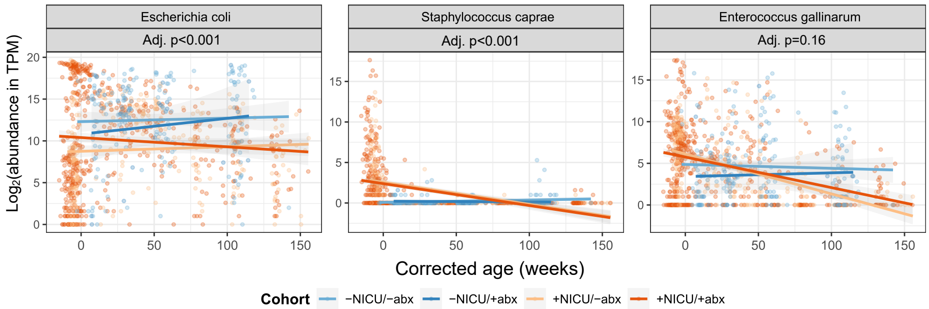
Escherichia coli
Adj. p<0.001
Staphylococcus caprae
Adj. p<0.001
Enterococcus gallinarum
Adj. p=0.16
Log2(abundance in TPM)
Corrected age (weeks)
Cohort
−NICU/−abx
−NICU/+abx
+NICU/−abx
+NICU/+abx

5b

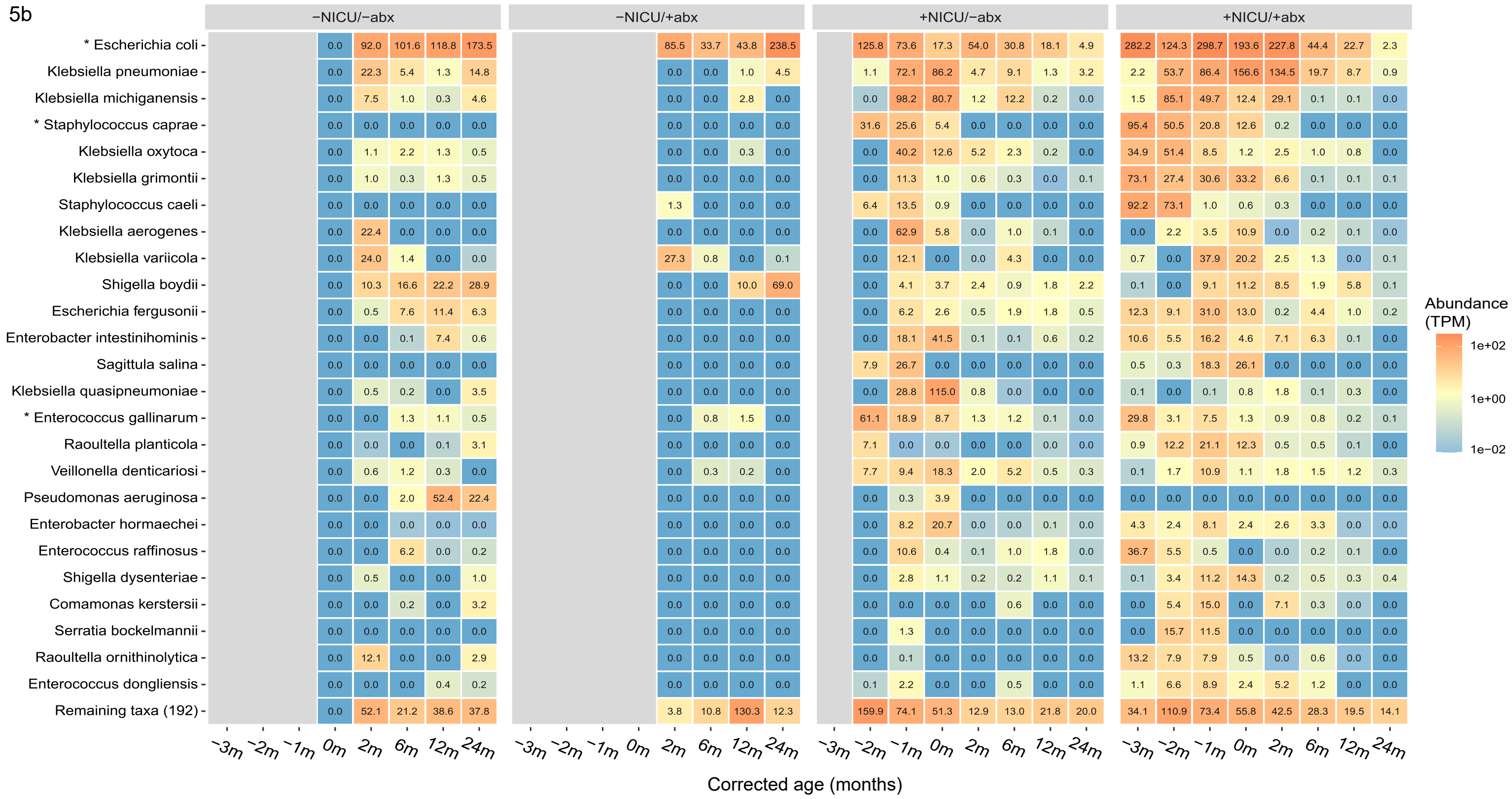
−NICU/−abx
−NICU/+abx
+NICU/−abx
+NICU/+abx
* Escherichia coli
Klebsiella pneumoniae
Klebsiella michiganensis
* Staphylococcus caprae
Klebsiella oxytoca
Klebsiella grimontii
Staphylococcus caeli
Klebsiella aerogenes
Klebsiella variicola
Shigella boydii
Escherichia fergusonii
Enterobacter intestinihominis
Sagittula salina
Klebsiella quasipneumoniae
* Enterococcus gallinarum
Raoultella planticola
Veillonella denticariosi
Pseudomonas aeruginosa
Enterobacter hormaechei
Enterococcus raffinosus
Shigella dysenteriae
Comamonas kerstersii
Serratia bockelmannii
Raoultella ornithinolytica
Enterococcus dongliensis
Remaining taxa (192)
−3m −2m −1m 0m 2m 6m 12m 24m
Corrected age (months)
Abundance (TPM)
1e+02
1e+00
1e−02

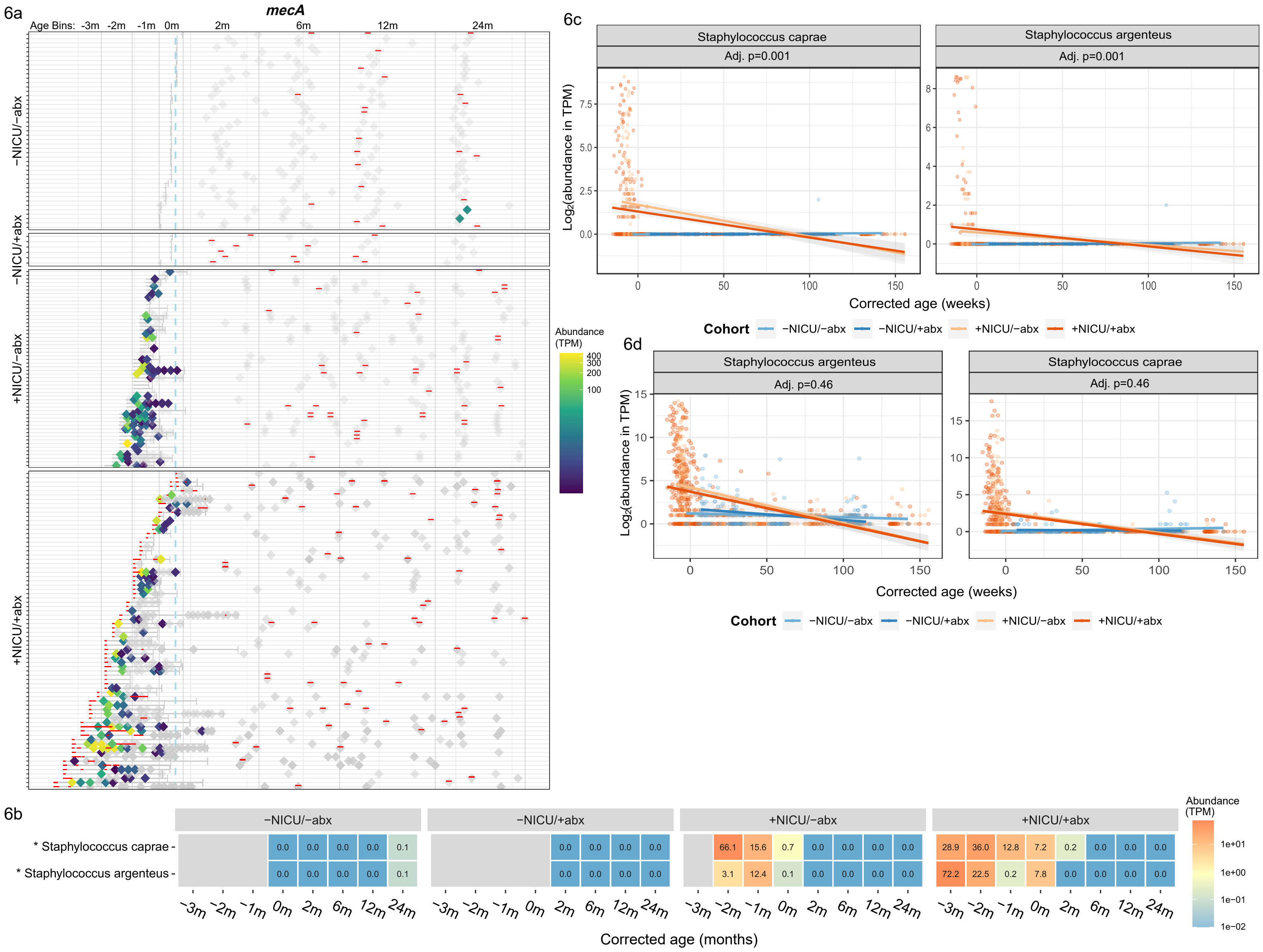
6a
mecA
Age Bins: -3m -2m -1m 0m 2m 6m 12m 24m
−NICU/−abx
−NICU/+abx
+NICU/−abx
+NICU/+abx
Abundance (TPM)
400
300
200
100
6c
Staphylococcus caprae
Adj. p=0.001
Staphylococcus argenteus
Adj. p=0.001
Log2(abundance in TPM)
Corrected age (weeks)
Cohort −NICU/−abx −NICU/+abx +NICU/−abx +NICU/+abx
6d
Staphylococcus argenteus
Adj. p=0.46
Staphylococcus caprae
Adj. p=0.46
Log2(abundance in TPM)
Corrected age (weeks)
Cohort −NICU/−abx −NICU/+abx +NICU/−abx +NICU/+abx
6b
−NICU/−abx
−NICU/+abx
+NICU/−abx
+NICU/+abx
* Staphylococcus caprae
* Staphylococcus argenteus
−NICU/−abx, * Staphylococcus caprae: 0.0 0.0 0.0 0.0 0.1
−NICU/−abx, * Staphylococcus argenteus: 0.0 0.0 0.0 0.0 0.1
−NICU/+abx, * Staphylococcus caprae: 0.0 0.0 0.0 0.0
−NICU/+abx, * Staphylococcus argenteus: 0.0 0.0 0.0 0.0
+NICU/−abx, * Staphylococcus caprae: 66.1 15.6 0.7 0.0 0.0 0.0 0.0
+NICU/−abx, * Staphylococcus argenteus: 3.1 12.4 0.1 0.0 0.0 0.0 0.0
+NICU/+abx, * Staphylococcus caprae: 28.9 36.0 12.8 7.2 0.2 0.0 0.0 0.0
+NICU/+abx, * Staphylococcus argenteus: 72.2 22.5 0.2 7.8 0.0 0.0 0.0 0.0
−3m −2m −1m 0m 2m 6m 12m 24m
Corrected age (months)
Abundance (TPM)
1e+01
1e+00
1e−01
1e−02

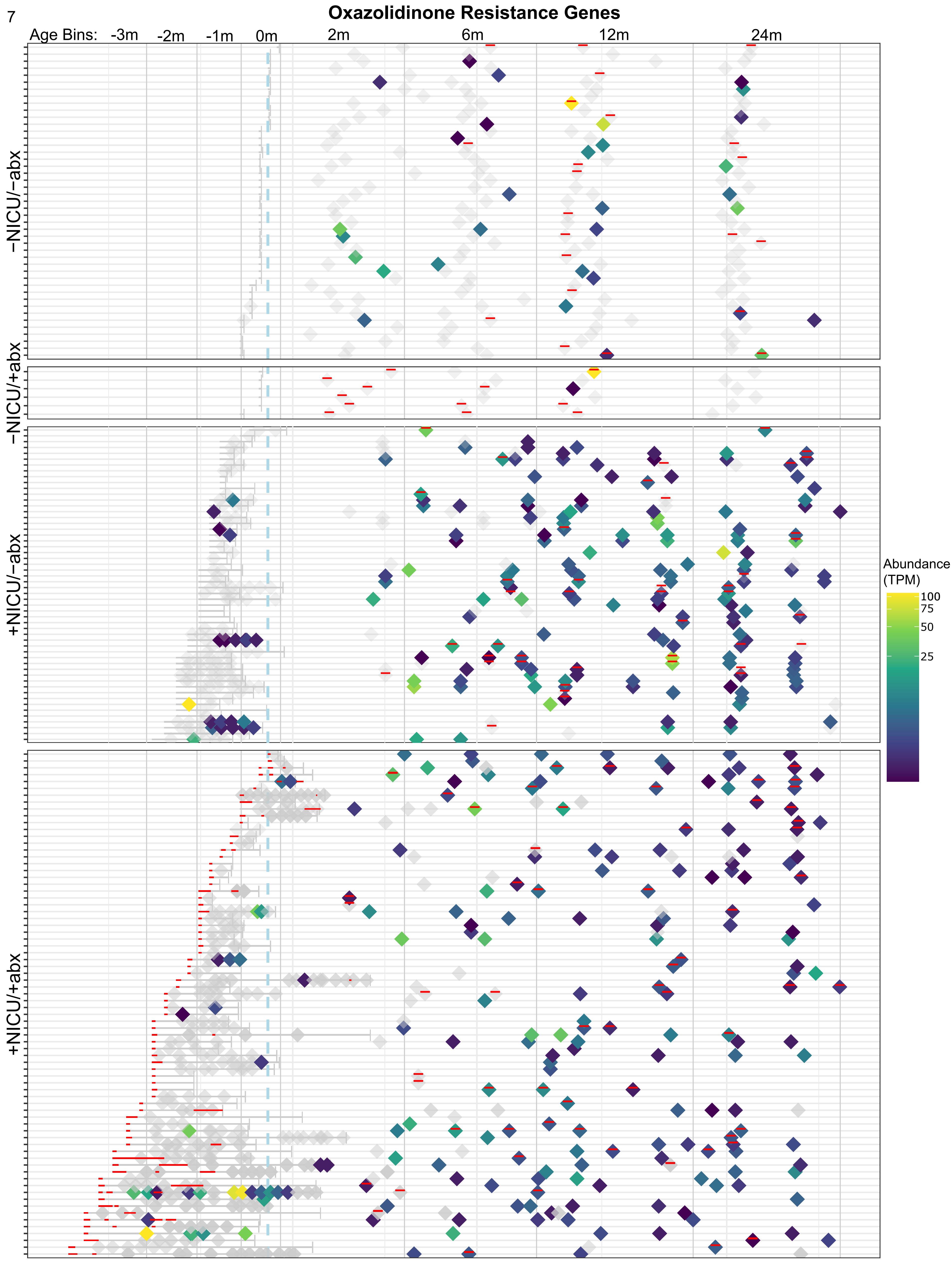

Oxazolidinone Resistance Genes
Age Bins:
-3m
-2m
-1m
0m
2m
6m
12m
24m
−NICU/−abx
−NICU/+abx
+NICU/−abx
+NICU/+abx
Abundance (TPM)
100
75
50
25

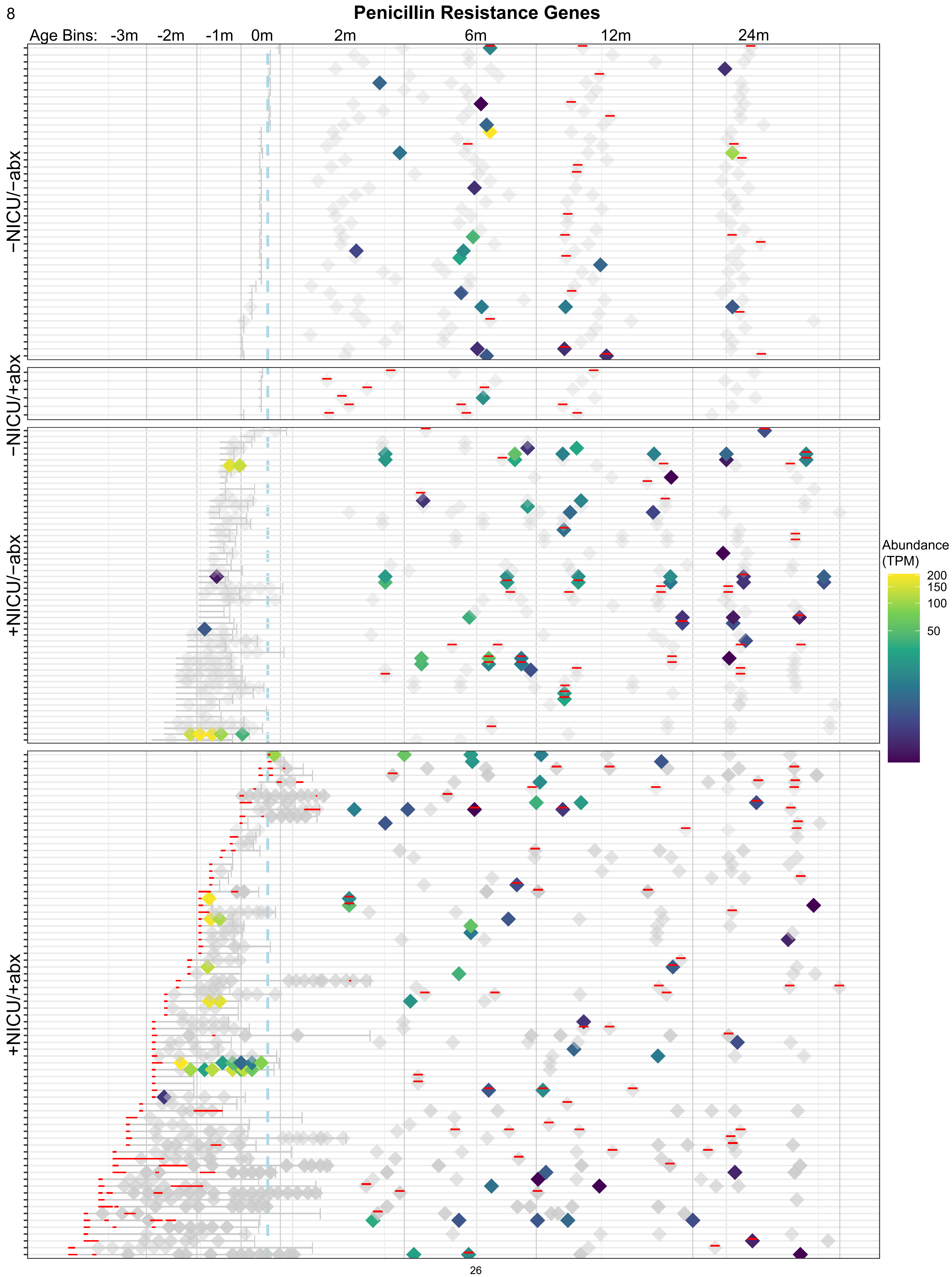
Penicillin Resistance Genes
Age Bins: -3m -2m -1m 0m 2m 6m 12m 24m
−NICU/−abx
−NICU/+abx
+NICU/−abx
+NICU/+abx
Abundance (TPM)
200
150
100
50

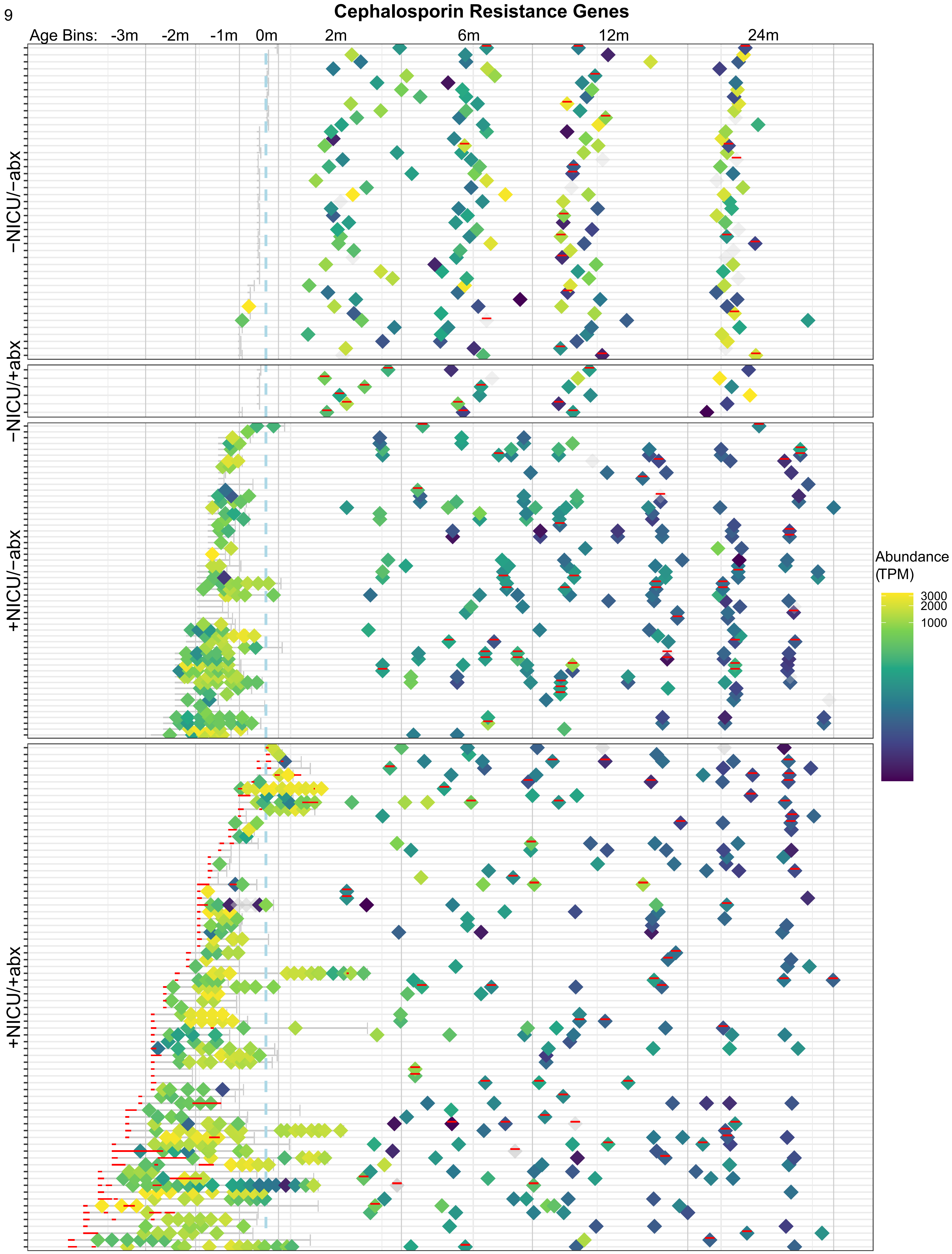
Cephalosporin Resistance Genes
Age Bins: -3m -2m -1m 0m 2m 6m 12m 24m
−NICU/−abx
−NICU/+abx
+NICU/−abx
+NICU/+abx
Abundance (TPM)
3000
2000
1000

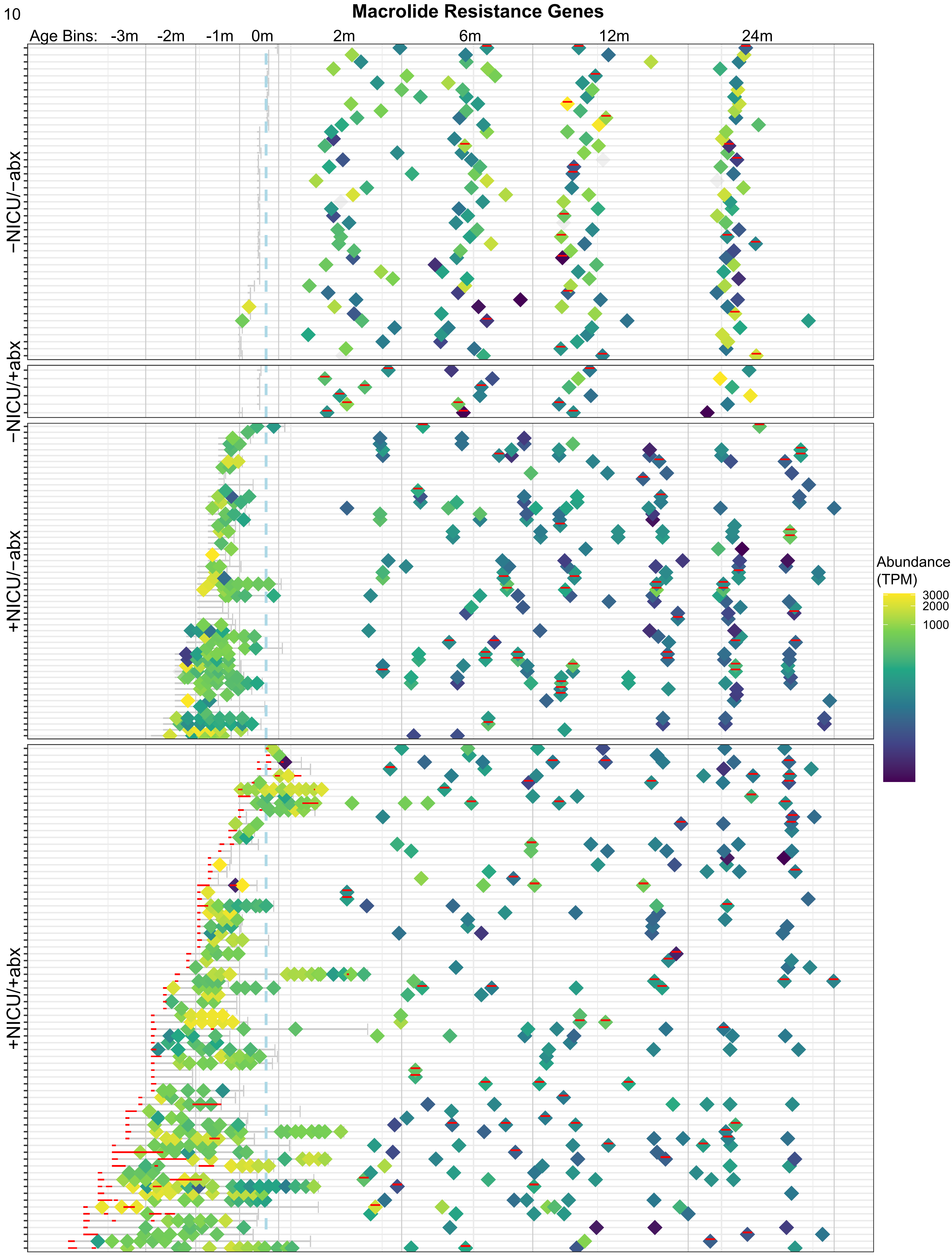
Macrolide Resistance Genes
Age Bins: -3m -2m -1m 0m 2m 6m 12m 24m
−NICU/−abx
−NICU/+abx
+NICU/−abx
+NICU/+abx
Abundance (TPM)
3000
2000
1000

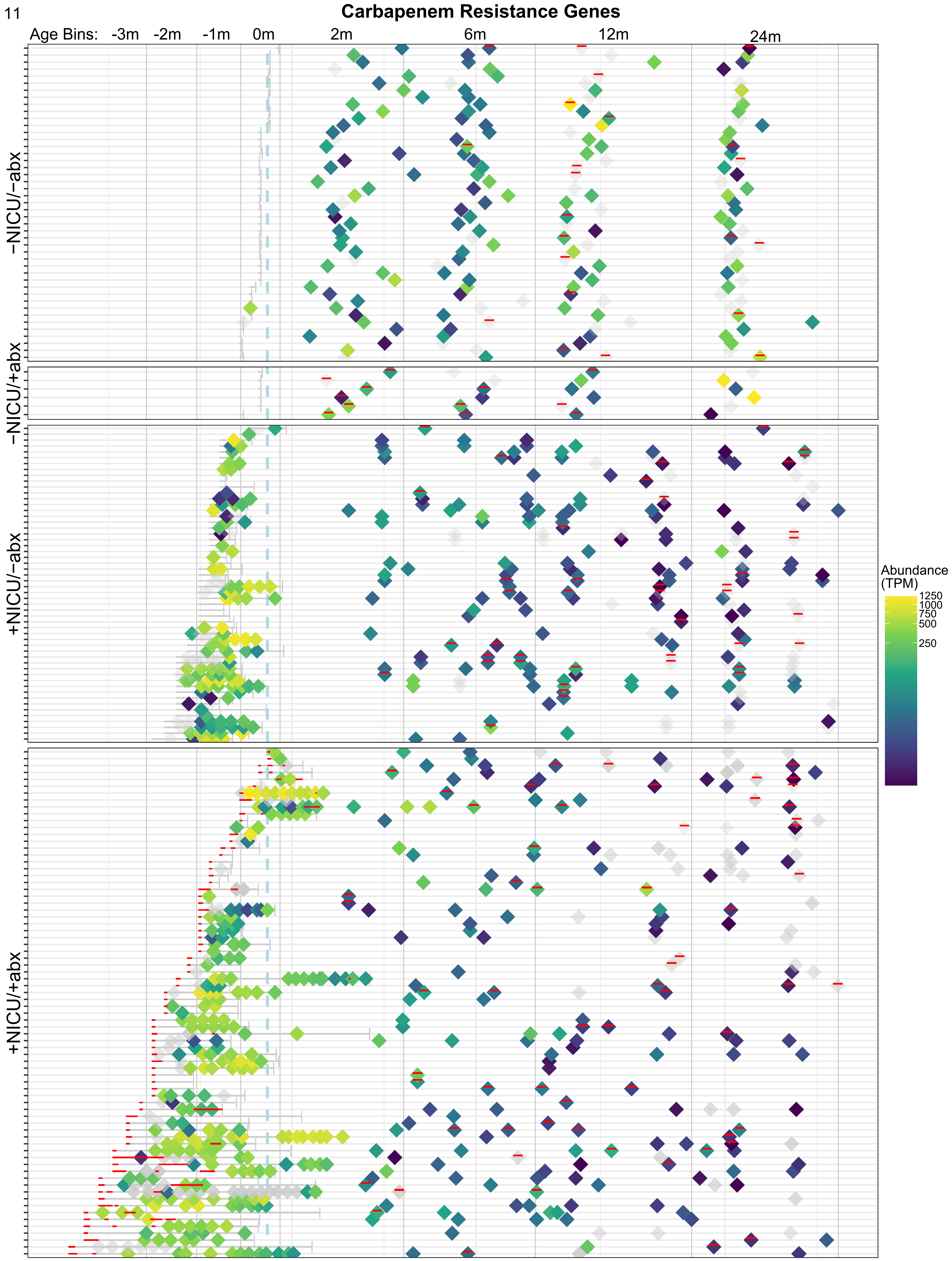
Carbapenem Resistance Genes
Age Bins: -3m -2m -1m 0m 2m 6m 12m 24m
−NICU/−abx
−NICU/+abx
+NICU/−abx
+NICU/+abx
Abundance (TPM)
1250
1000
750
500
250

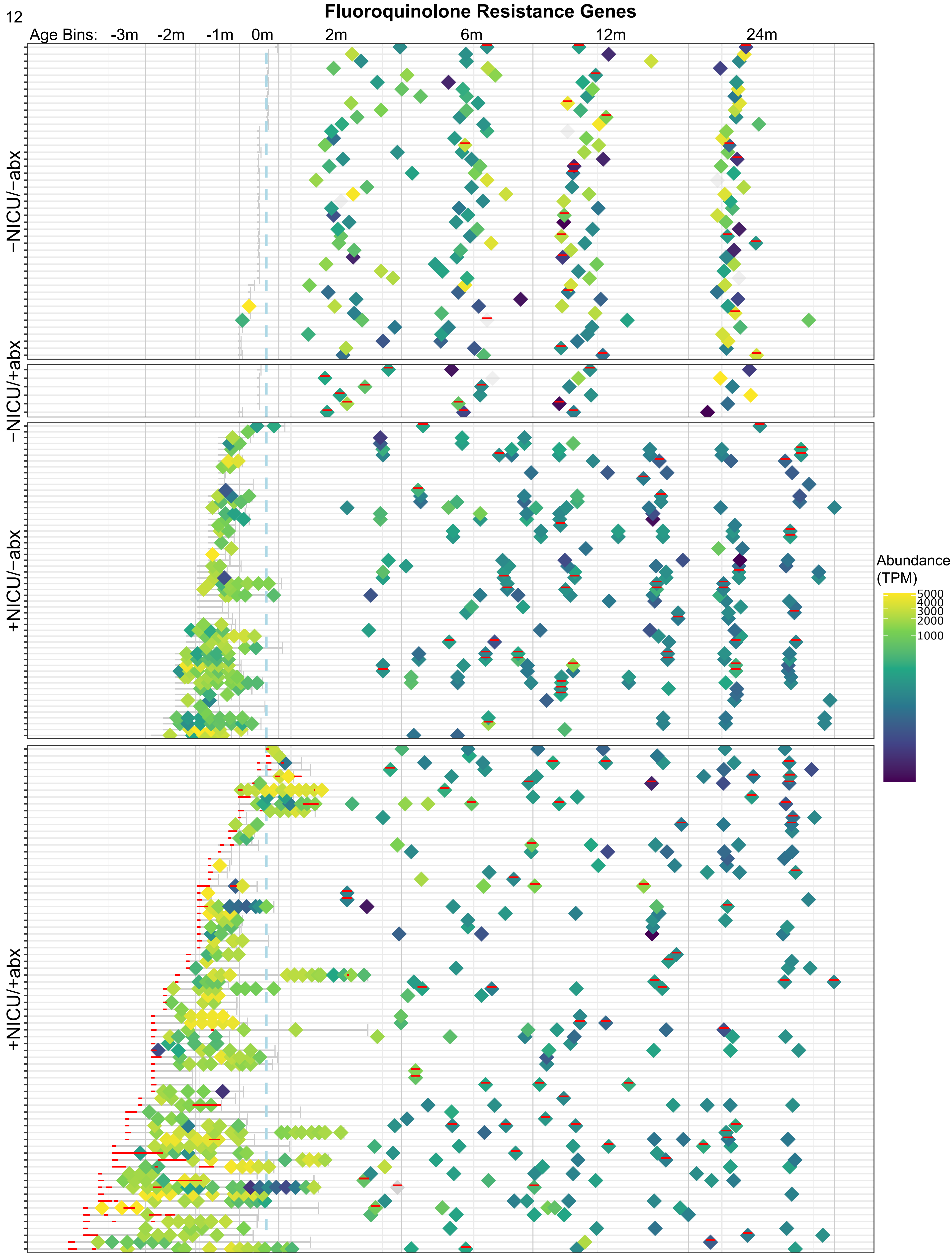

Fluoroquinolone Resistance Genes
Age Bins: -3m -2m -1m 0m 2m 6m 12m 24m
−NICU/−abx
−NICU/+abx
+NICU/−abx
+NICU/+abx
Abundance (TPM)
5000
4000
3000
2000
1000

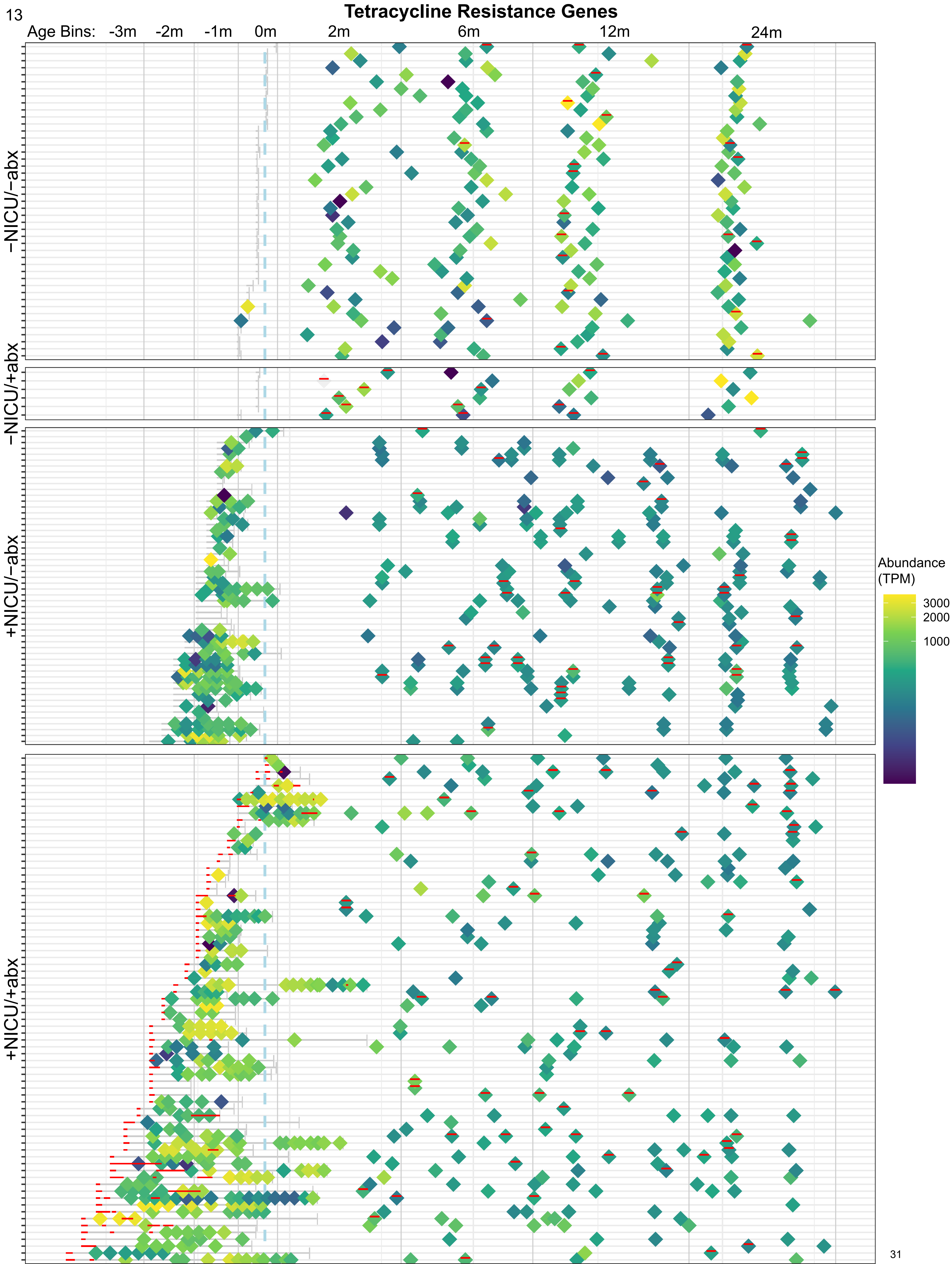
Tetracycline Resistance Genes
Age Bins:
-3m
-2m
-1m
0m
2m
6m
12m
24m
−NICU/−abx
−NICU/+abx
+NICU/−abx
+NICU/+abx
Abundance (TPM)
3000
2000
1000

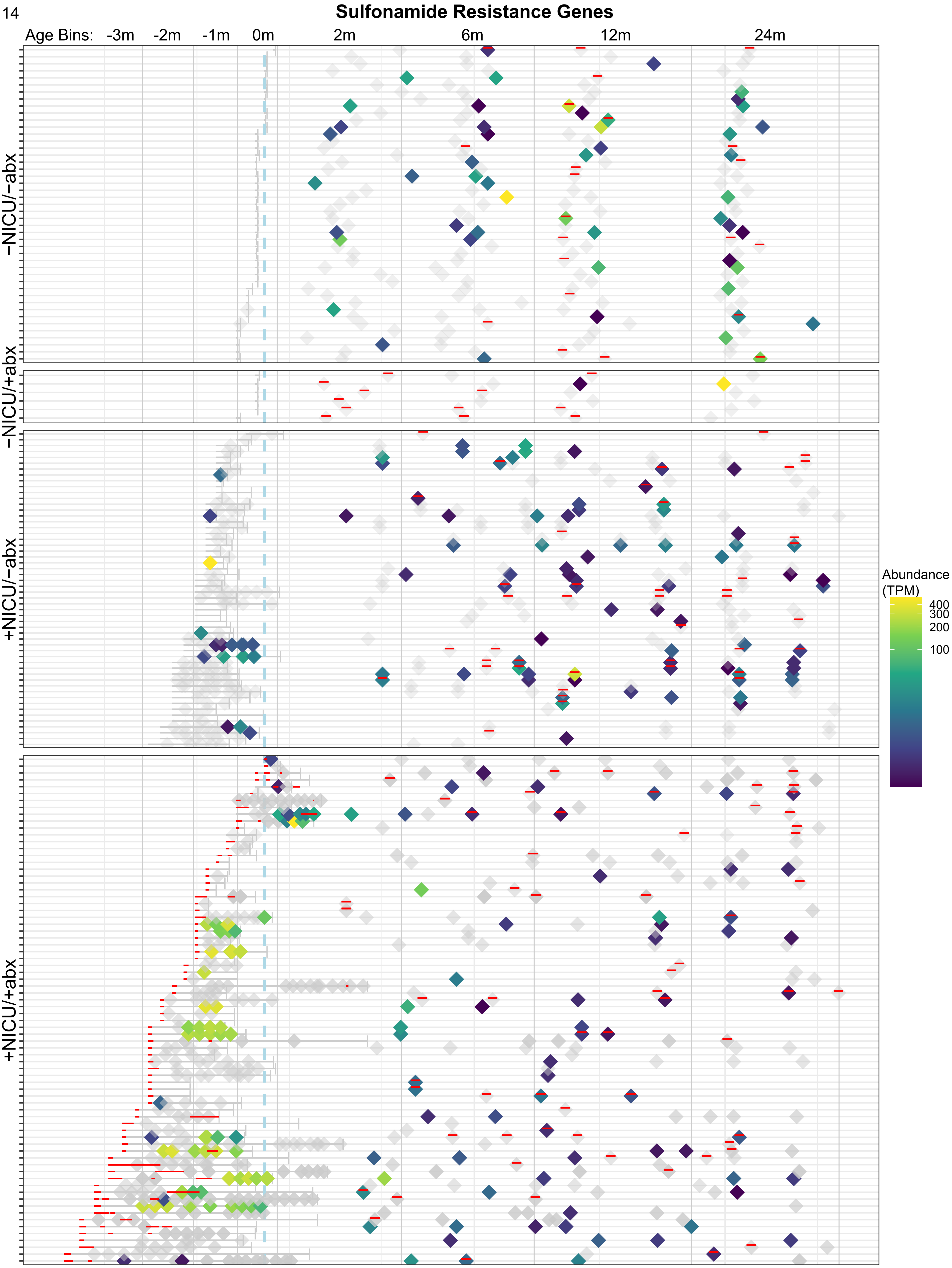
Sulfonamide Resistance Genes
Age Bins: -3m -2m -1m 0m 2m 6m 12m 24m
−NICU/−abx
−NICU/+abx
+NICU/−abx
+NICU/+abx
Abundance (TPM)
400
300
200
100

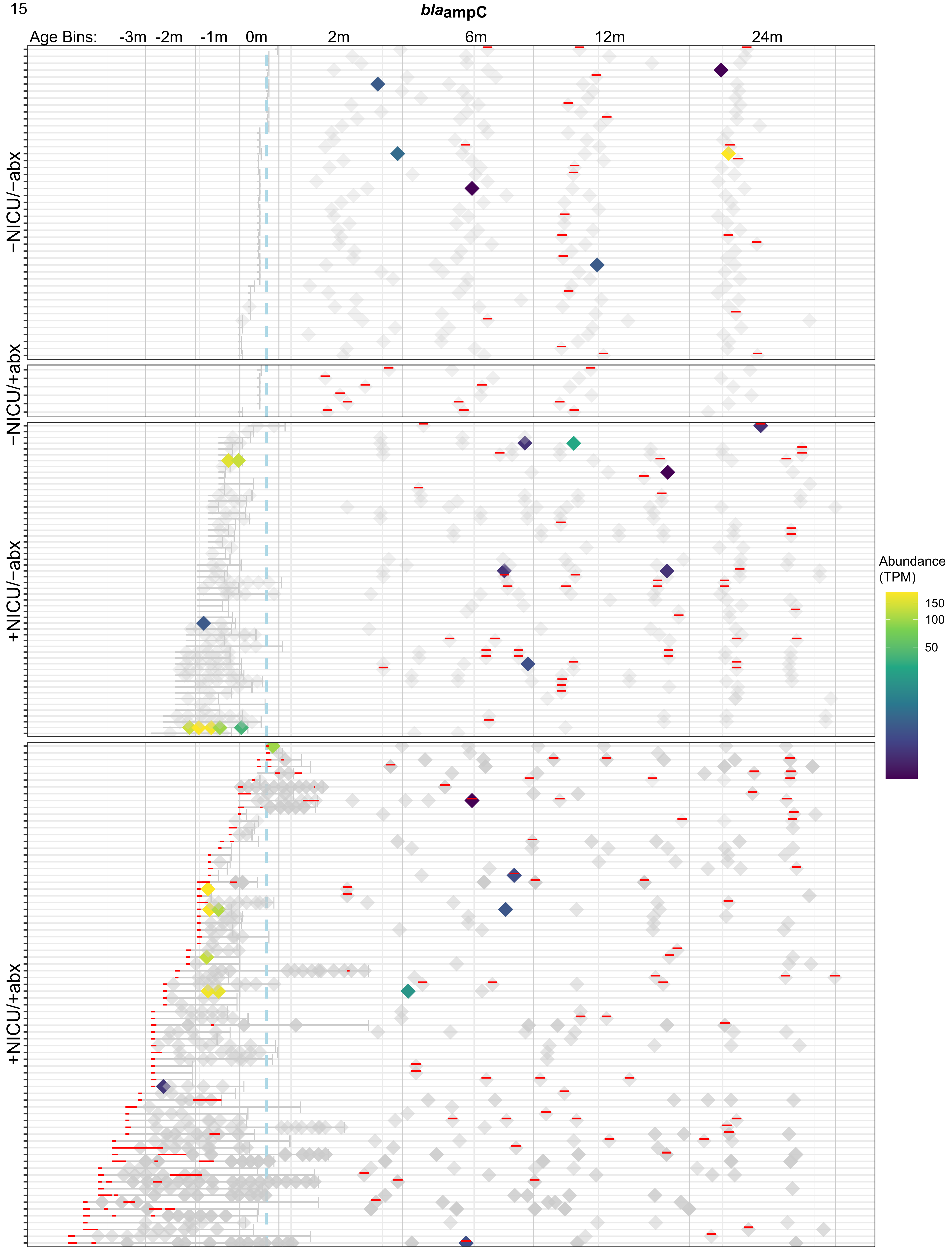

*bla*ampC
Age Bins: -3m -2m -1m 0m 2m 6m 12m 24m
−NICU/−abx
−NICU/+abx
+NICU/−abx
+NICU/+abx
Abundance (TPM)
150
100
50

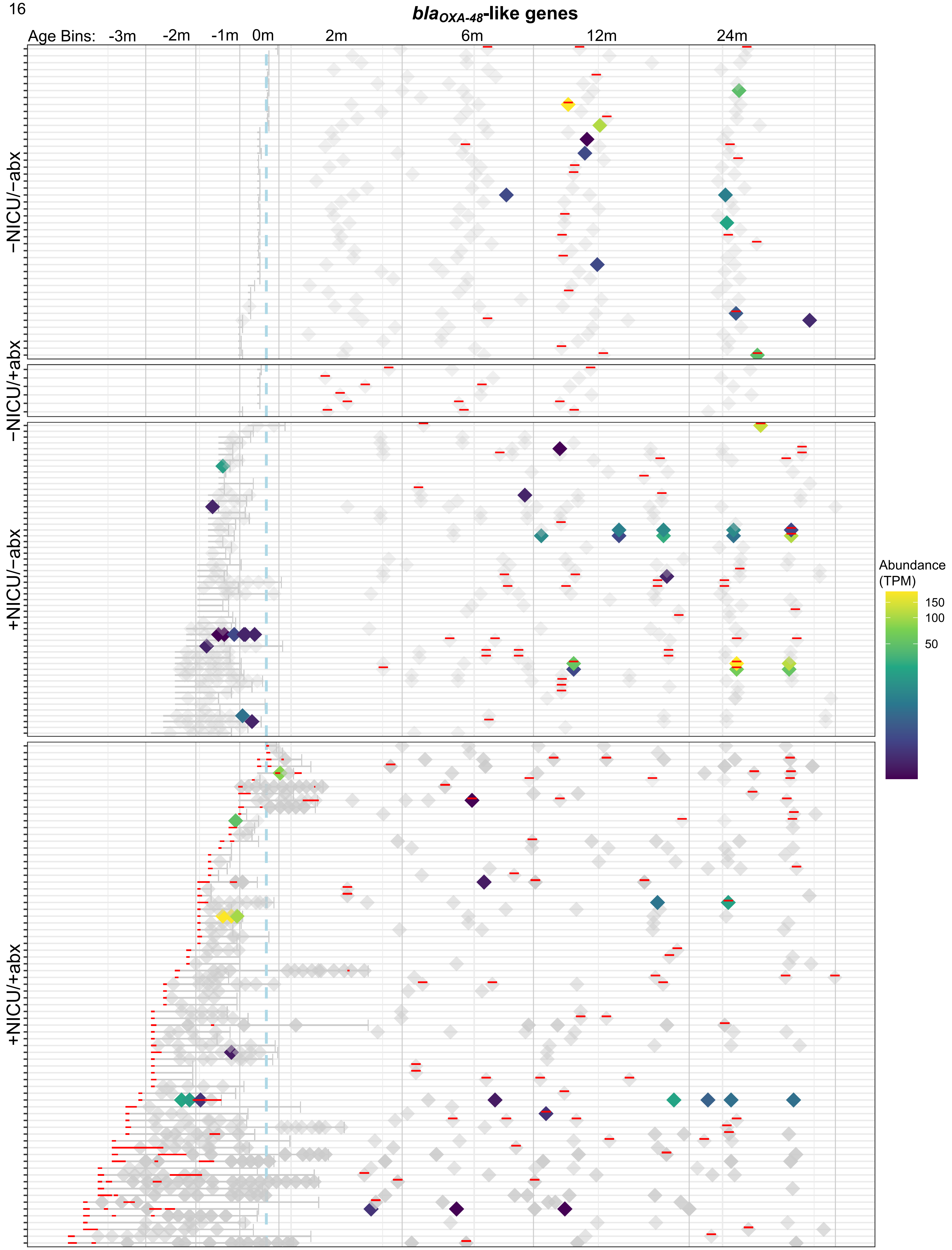

blaOXA-48-like genes
Age Bins:
-3m
-2m
-1m
0m
2m
6m
12m
24m
−NICU/−abx
−NICU/+abx
+NICU/−abx
+NICU/+abx
Abundance
(TPM)
150
100
50

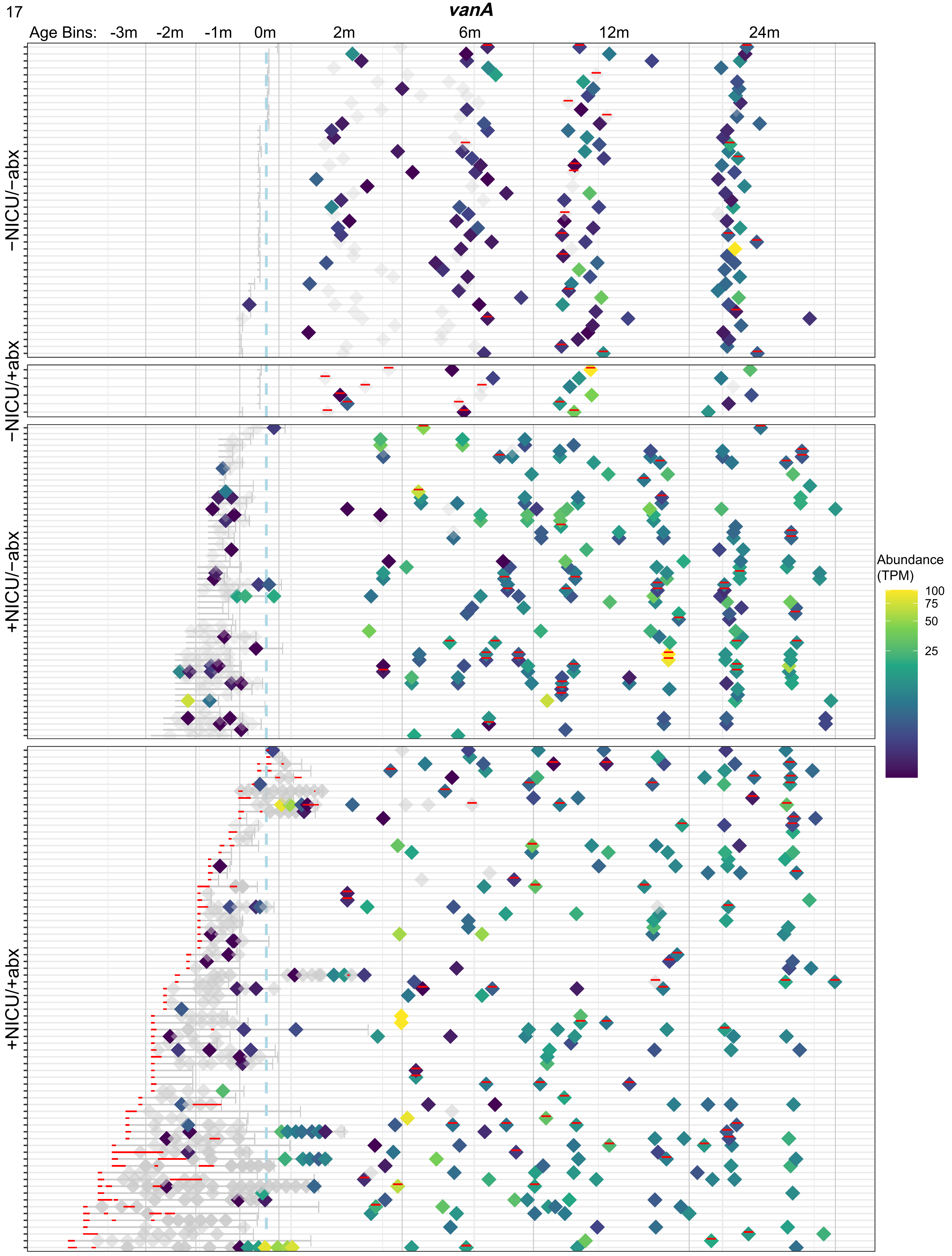
vanA
Age Bins: -3m -2m -1m 0m 2m 6m 12m 24m
−NICU/−abx
−NICU/+abx
+NICU/−abx
+NICU/+abx
Abundance (TPM)
100
75
50
25

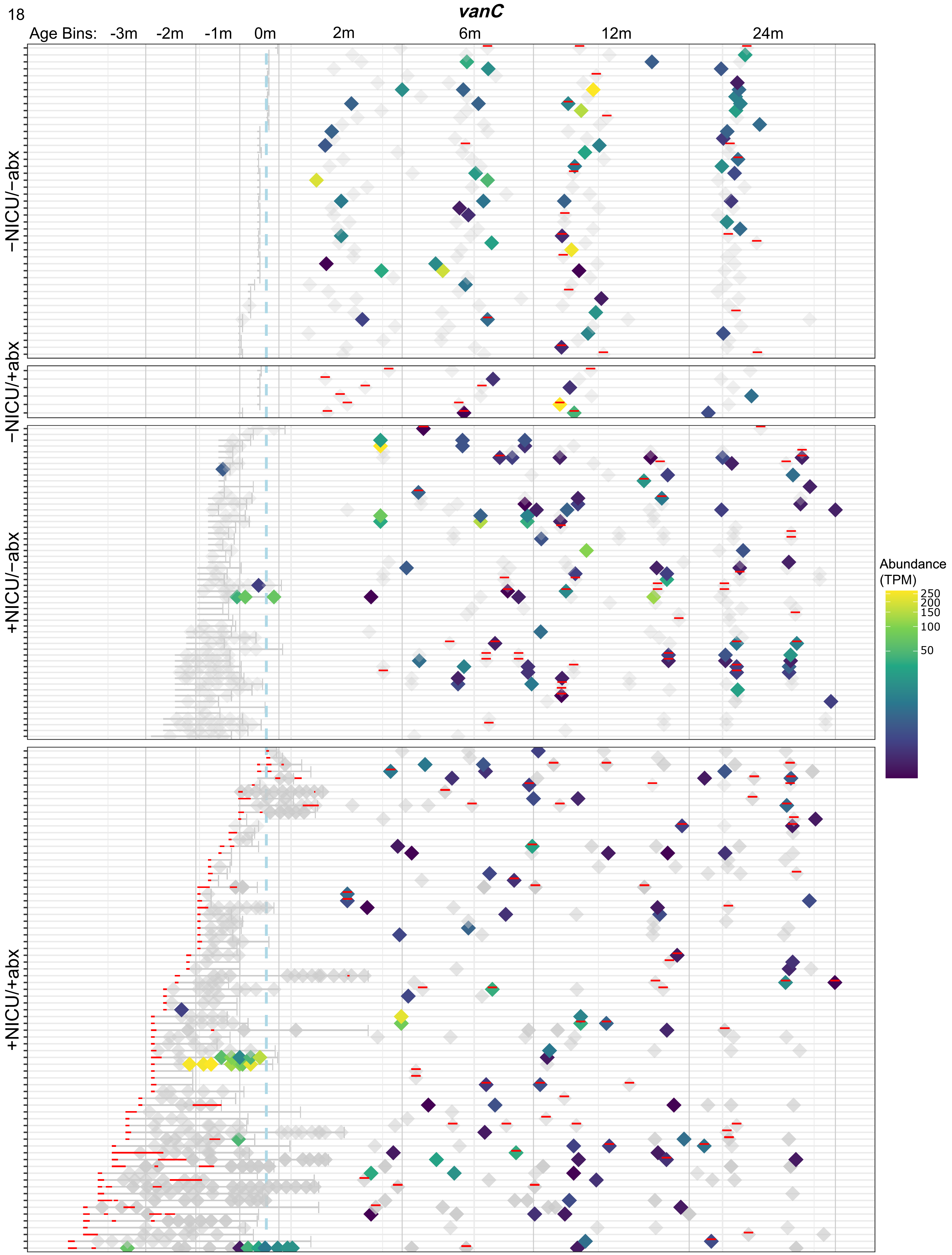
vanC
Age Bins: -3m -2m -1m 0m 2m 6m 12m 24m
−NICU/−abx
−NICU/+abx
+NICU/−abx
+NICU/+abx
Abundance (TPM)
250
200
150
100
50

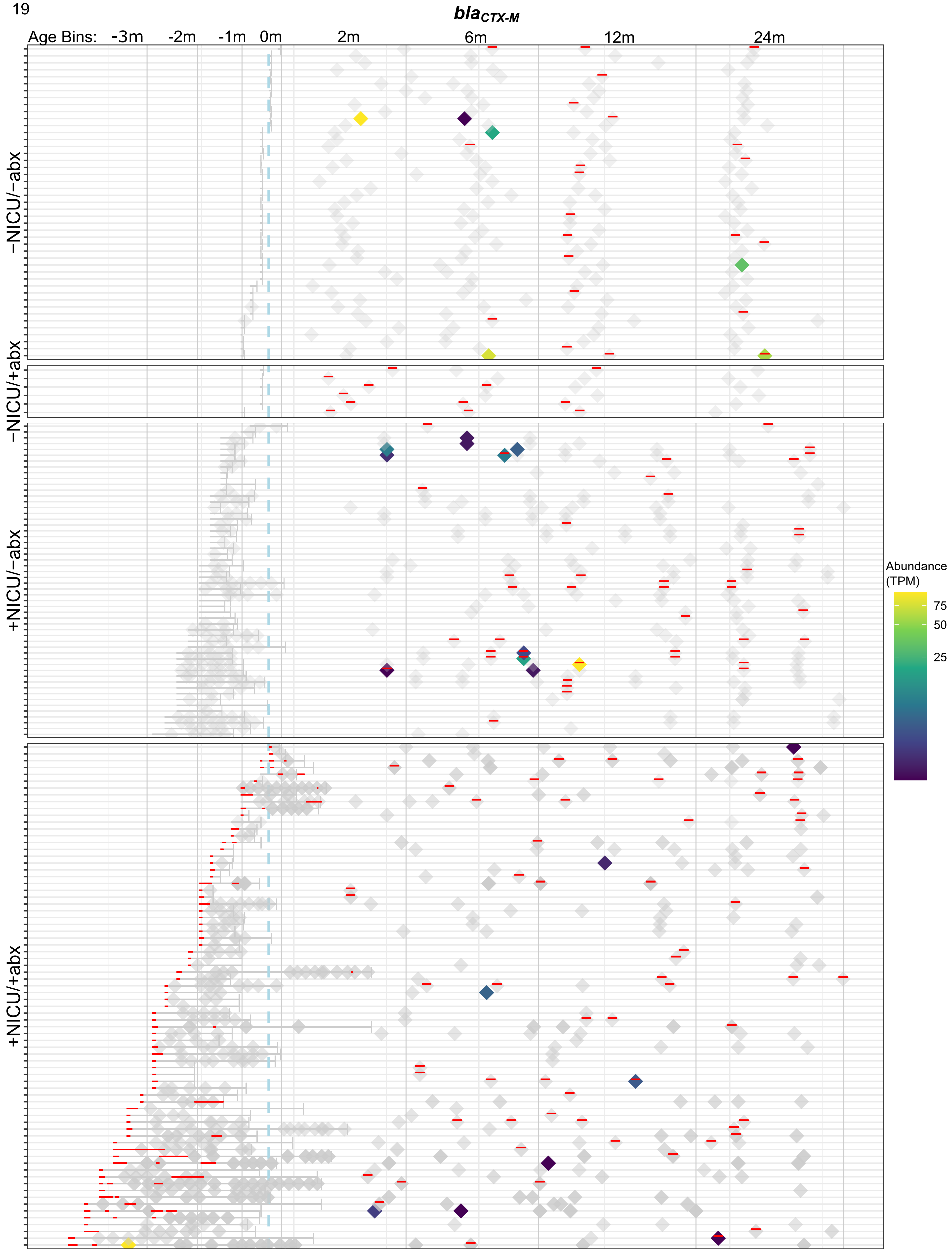
blaCTX-M
Age Bins: -3m -2m -1m 0m 2m 6m 12m 24m
−NICU/−abx
−NICU/+abx
+NICU/−abx
+NICU/+abx
Abundance (TPM)
75
50
25

20

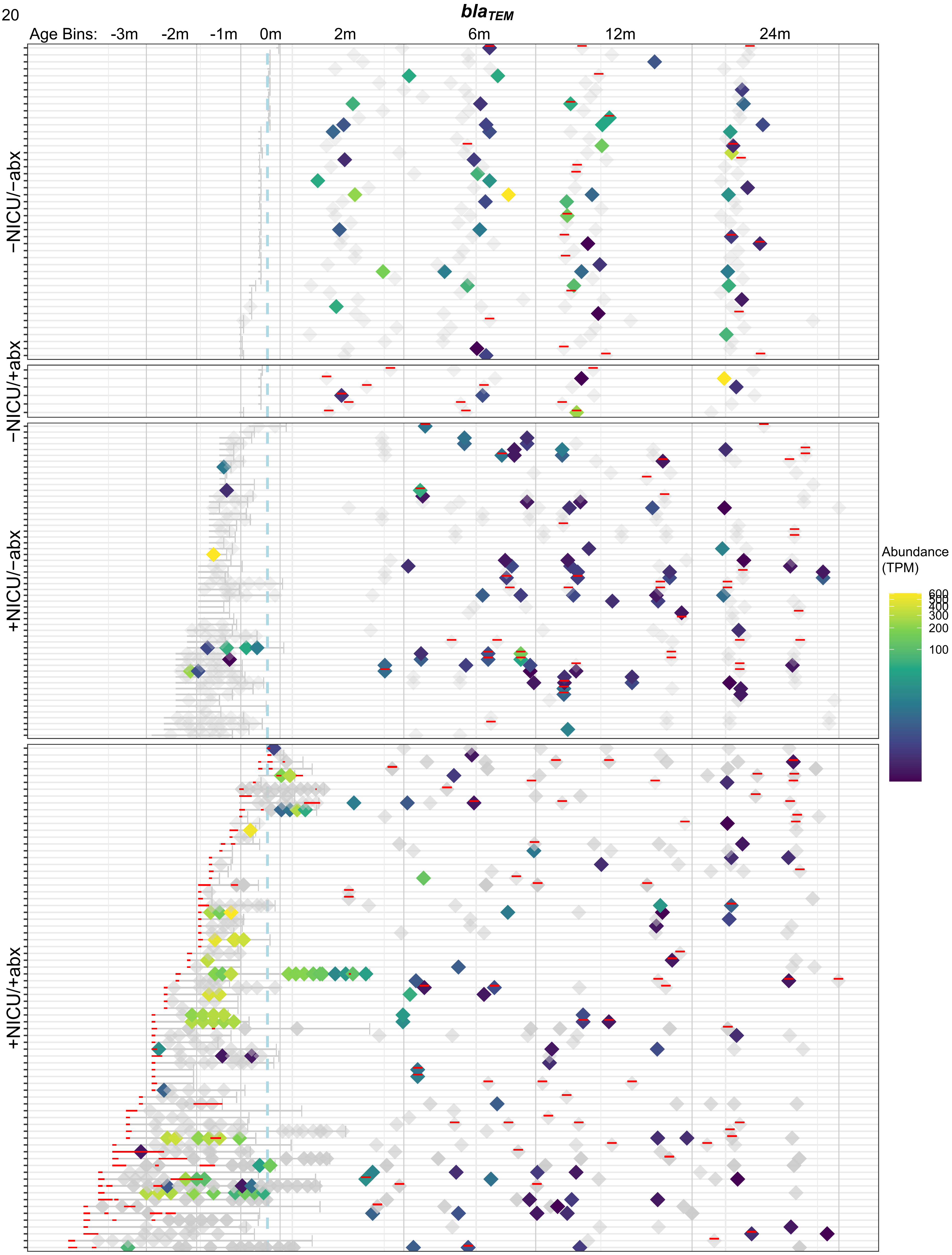

blaTEM
Age Bins: -3m -2m -1m 0m 2m 6m 12m 24m
−NICU/−abx
−NICU/+abx
+NICU/−abx
+NICU/+abx
Abundance (TPM)
600
500
400
300
200
100

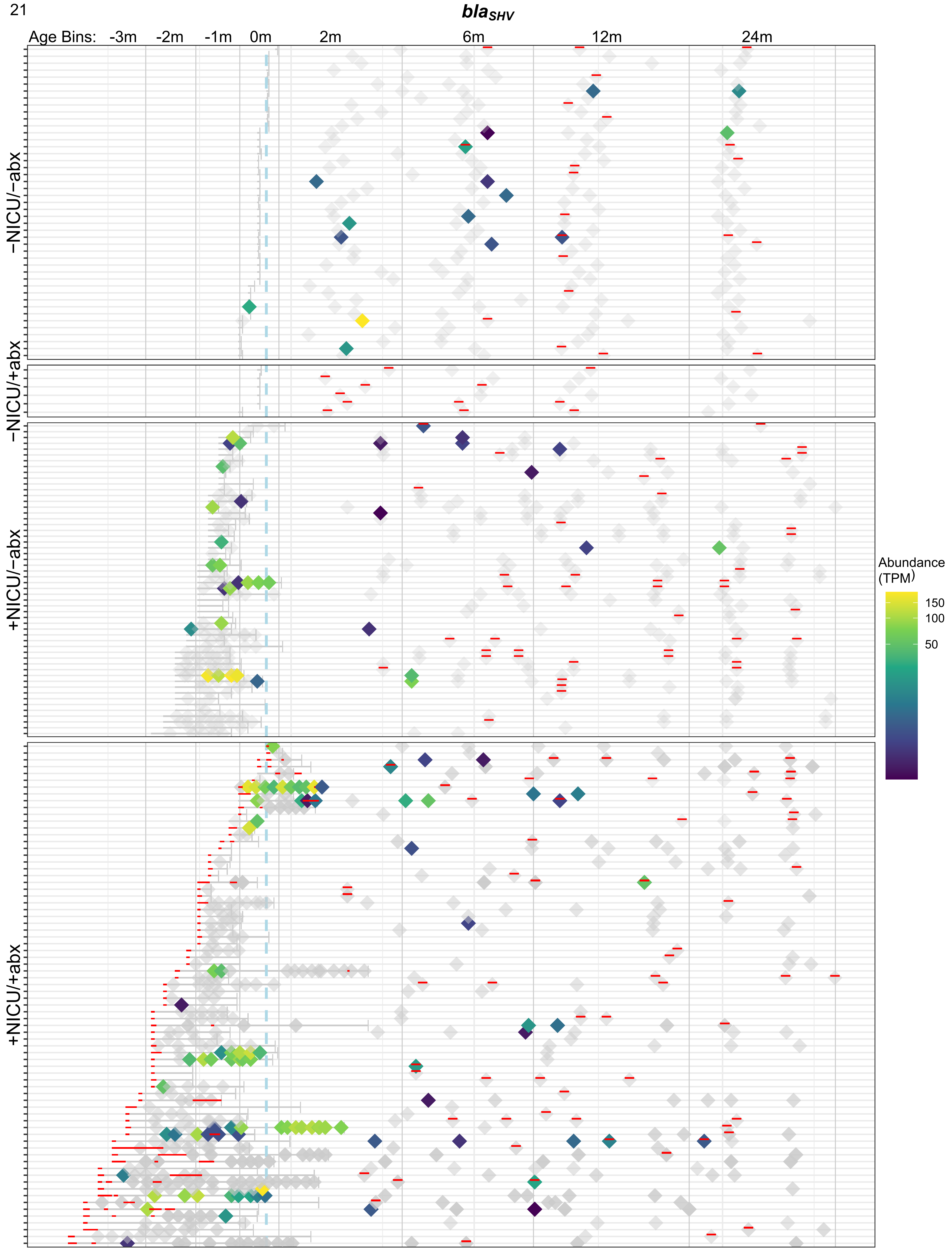

bla_SHV
Age Bins:
-3m
-2m
-1m
0m
2m
6m
12m
24m
−NICU/−abx
−NICU/+abx
+NICU/−abx
+NICU/+abx
Abundance (TPM)
150
100
50

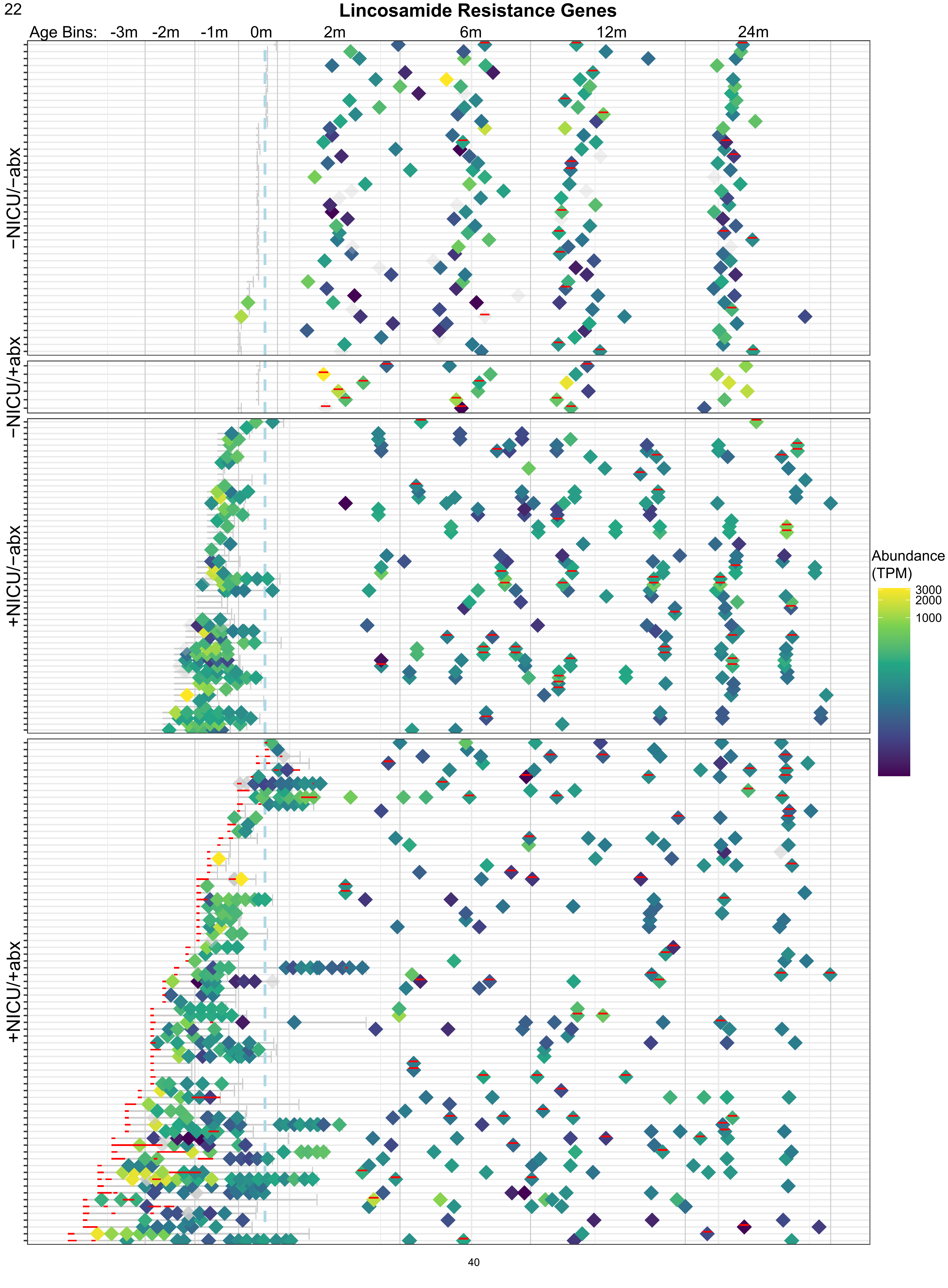
Lincosamide Resistance Genes
Age Bins: -3m -2m -1m 0m 2m 6m 12m 24m
−NICU/−abx
−NICU/+abx
+NICU/−abx
+NICU/+abx
Abundance (TPM)
3000
2000
1000

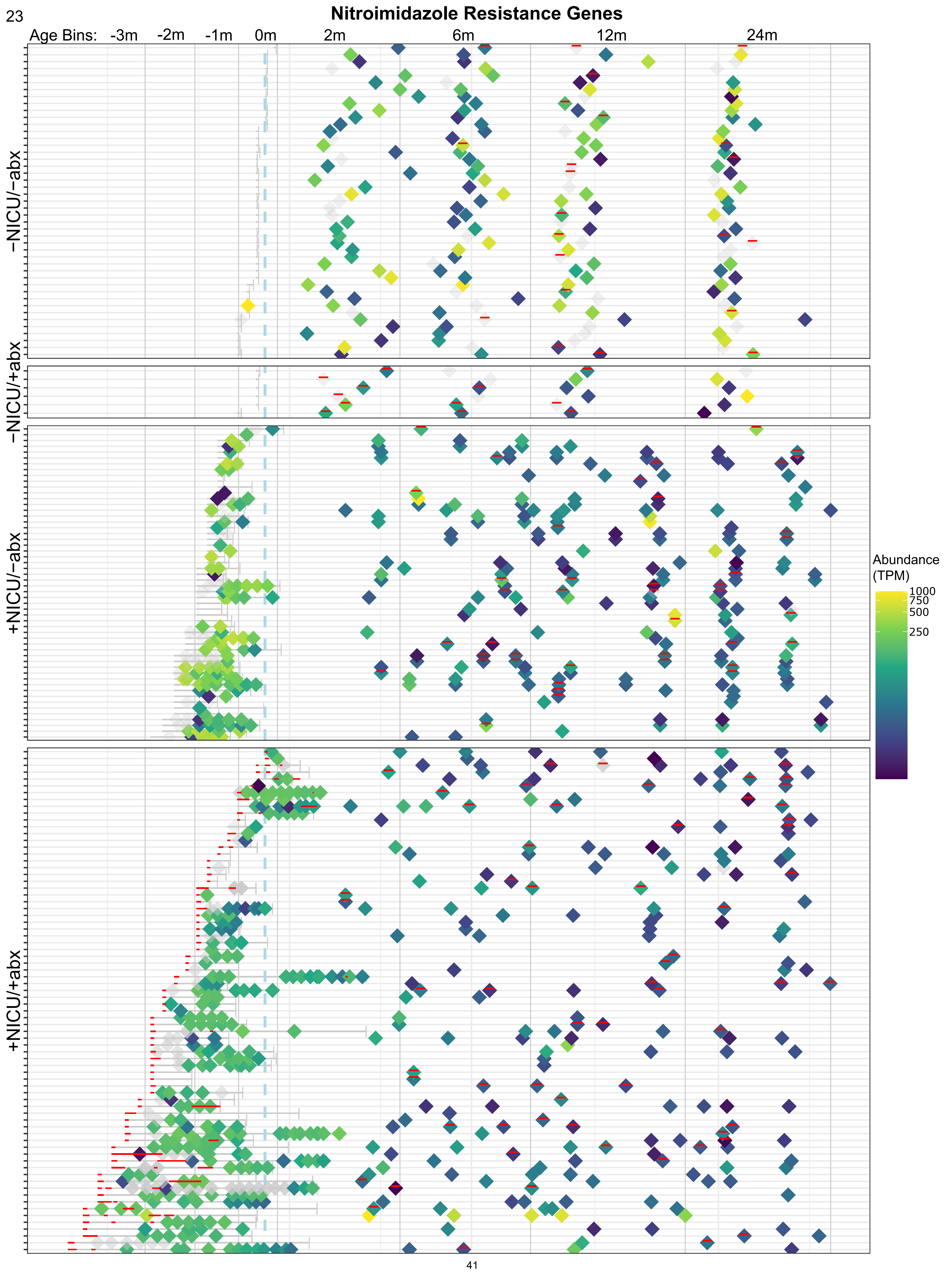

Nitroimidazole Resistance Genes
Age Bins: -3m -2m -1m 0m 2m 6m 12m 24m
−NICU/−abx
−NICU/+abx
+NICU/−abx
+NICU/+abx
Abundance (TPM)
1000
750
500
250

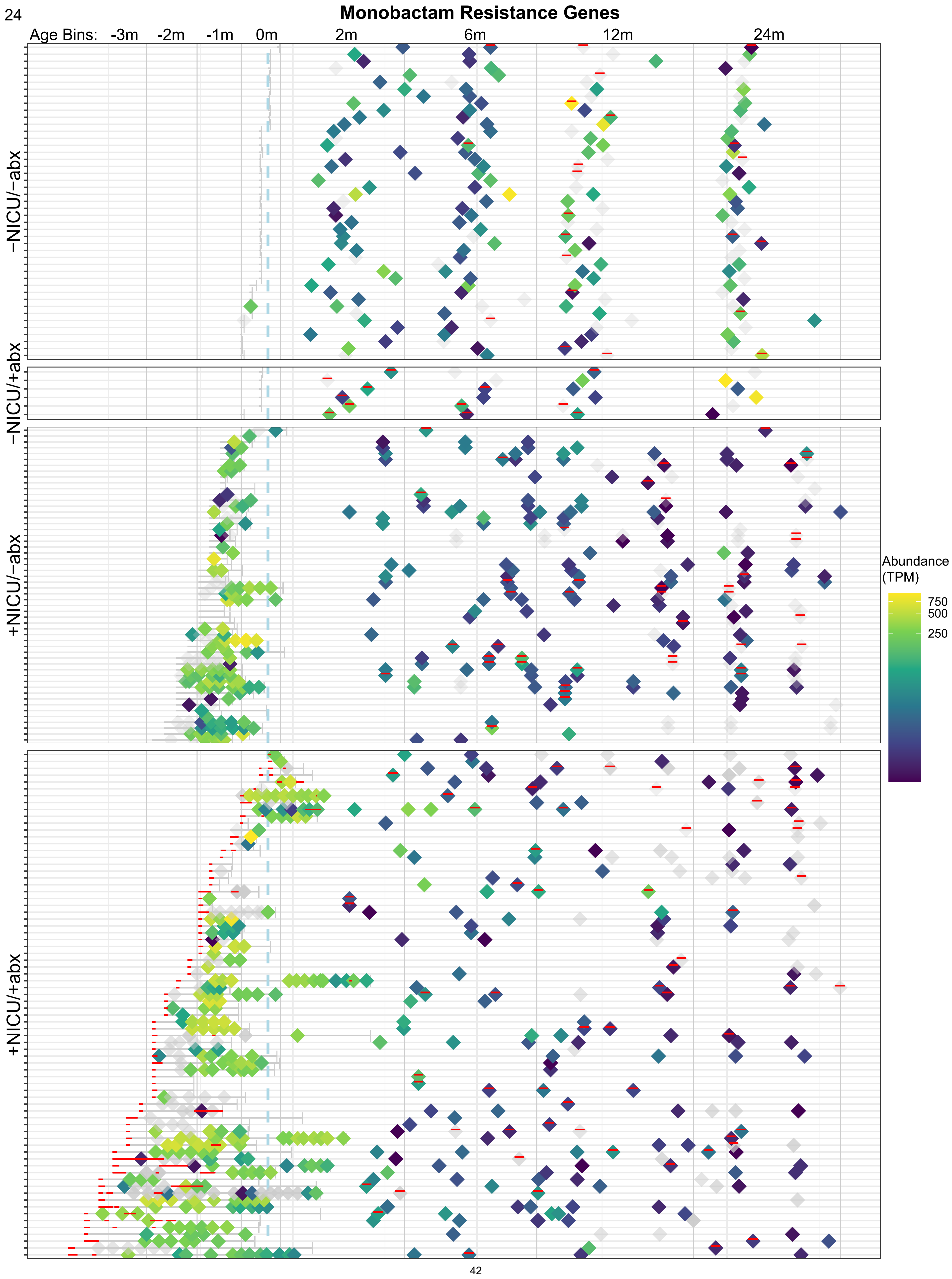
Monobactam Resistance Genes
Age Bins:
-3m
-2m
-1m
0m
2m
6m
12m
24m
−NICU/−abx
−NICU/+abx
+NICU/−abx
+NICU/+abx
Abundance
(TPM)
750
500
250
